# Post COVID-19 Condition, Work Ability and Occupational Changes: Results from a Population-based Cohort

**DOI:** 10.1101/2023.04.17.23288664

**Authors:** Philipp Kerksieck, Tala Ballouz, Sarah R. Haile, Celine Schumacher, Joanne Lacy, Anja Domenghino, Jan S. Fehr, Georg F. Bauer, Holger Dressel, Milo A. Puhan, Dominik Menges

**Affiliations:** Epidemiology, Biostatistics and Prevention Institute (EBPI), University of Zurich (UZH), Zurich, Switzerland; Department of Visceral and Transplantation Surgery, University Hospital Zurich (USZ), Zurich, Switzerland

## Abstract

**Background:** Evidence from population-based studies on the impact of post COVID-19 condition (PCC) on ability to work is limited but critical due to its high prevalence among individuals of working-age.

**Objective:** To evaluate the association between PCC, work ability, and occupational changes.

**Design:** Population-based, longitudinal cohort.

**Setting:** General population, Canton of Zurich, Switzerland.

**Participants:** 672 adults of working-age with SARS-CoV-2 infection.

**Measurements:** Current work ability, work ability related to physical and mental demands, and estimated future work ability in 2 years (assessed using Work Ability Index), as well as PCC-related occupational changes at one year after infection.

**Results:** There was very strong evidence that current work ability scores were 0.62 (95% confidence interval (CI) 0.30 to 0.95) points lower among those with PCC compared to those without. Similarly, there was very strong evidence for lower odds of reporting higher work ability with respect to physical (odds ratio (OR) 0.30, 95% CI 0.20 to 0.46) and mental (OR 0.40, 0.27 to 0.62) demands among those with PCC compared to those without. Higher age and history of psychiatric diagnosis were associated with a more substantial reduction in current work ability. 5.8% of those with PCC reported direct effects of PCC on their occupational situation, with 1.6% of those with PCC completely dropping out of the workforce and 43% of those with PCC-related occupational changes reporting financial difficulties as a result.

**Limitations:** Selection, use of self-reported outcome measures, and limited generalizability to individuals with most severe COVID-19 or following vaccination.

**Conclusions:** These findings highlight the need for providing support and interdisciplinary interventions to individuals affected by PCC to help them maintain or regain their work ability and productivity.

**Primary Funding Source:** Federal Office of Public Health, Department of Health of the Canton of Zurich, University of Zurich Foundation, Switzerland.

**Study Registration:** ISRCTN14990068.

## Introduction

Post COVID-19 condition (PCC) affects 10-20% of individuals infected with SARS-CoV-2 (1–11). Symptoms associated with PCC are varied and can be physical (e.g., fatigue, post-exertional malaise, pain, and dyspnea) or mental (most commonly memory and concentration difficulties) (2,12,13). Many of these symptoms adversely impact individuals’ everyday functioning, including impairments to their ability to engage in physical activities and participate in social life and work (1,2,14). The prevalence of PCC is highest among those of working age (11,12) and the resulting socioeconomic implications are likely considerable (15,16). While it is important to develop effective management strategies and interventions to reduce the health burden of PCC, it is thus critical to also consider its impact on the workforce and establish sensible pathways to restore occupational participation in those severely affected.

Few studies have evaluated the association of PCC with work-related functioning or subsequent occupational changes (17–27). Most focused on describing work absenteeism and found that 11%-50% of workers do not return to work several months after COVID-19 (2,14). Various individual, organizational, and systemic aspects (e.g., supportive return-to-work policies) contribute to successful return to work after an illness, including having sufficient actual work ability (28–30). Work ability is a multifactorial measure frequently used in clinical practice and research to assess the degree to which an individual is physically and mentally able to cope with demands at work (31–33). In addition to short- and long-term sickness absence (34–37), poor work ability is also associated with early retirement (38,39) and disability at work (39,40), all of which carry large repercussions for the labor market and economy. Rehabilitation programs targeted at the working-age population generally aim to improve or preserve work ability. Given the substantial prevalence of PCC and its potential for long-term work-related consequences, understanding the association of PCC with work ability is crucial for the development of policies and multidisciplinary strategies aimed at supporting affected individuals in their recovery.

In this study, we aimed to comprehensively evaluate the association between PCC, work ability, and occupational changes in a working-age population within a prospective population-based cohort of SARS-CoV-2 infected individuals.

## Methods

### Study Design and Participants

We used data from a population-based, prospective, observational cohort of individuals with diagnosed SARS-CoV-2 infection from the Canton of Zurich, Switzerland (Zurich SARS-CoV-2 Cohort; ISRCTN14990068) (41). Based on mandatory reporting of all SARS-CoV-2 infections to the Department of Health of the Canton of Zurich, we prospectively invited on a daily basis an age-stratified (18–39 years, 40–64 years, ≥65 years), random sample of eligible individuals diagnosed between 06 August 2020 and 19 January 2021 for study participation. Eligibility criteria were being 18 years or older, able to follow study procedures, residing in the Canton of Zurich, and having sufficient knowledge of the German language. All participants were enrolled upon or shortly after diagnosis, infected with wildtype SARS-CoV-2, and were unvaccinated at time of infection. In this study we included individuals of working age (18–64 years old; the retirement age is 65 years in Switzerland (42)) who did not report being retired at enrollment. To ensure that evaluated outcomes were not related to reinfection with SARS-CoV-2 over the course of follow-up, we excluded individuals reporting a reinfection event. The study was approved by the ethics committee of the Canton of Zurich (BASEC-Nr. 2020-01739) and we obtained written or electronic consent from all participants.

### Data Sources

We collected data using electronic questionnaires. At baseline immediately after enrollment, we collected data on the acute primary infection (i.e., symptoms, severity), pre-existing comorbidities (any of hypertension, diabetes, cardiovascular disease, chronic respiratory disease, chronic kidney disease, malignancy, or immune suppression), pre-infection health status, and socio-demographic characteristics. In this ongoing cohort, we collect follow-up data on participants’ health trajectories in regular intervals after infection (9,10). At 12 months, we additionally elicited measures of work ability and asked participants to report any occupational changes over the first 12 months post-infection. Simultaneously, we asked participants to report any pre-existing psychiatric diagnoses before infection and any new or worsened psychiatric diagnoses during follow-up. Participants were also asked to provide further details in free text fields. One researcher (DM) additionally conducted personal phone interviews with participants for whom questionnaire information was not unequivocal (n=4).

### Outcome Measurement

We assessed self-perceived work ability using selected measures from the Work Ability Index, a validated and frequently used instrument for assessing work ability (31,32,34). The primary outcome was the current work ability scale (score from 1–10, with 10 being best ability and 0 no ability to work). In sensitivity analyses, we categorized current work ability into poor (scores ≤6), moderate (scores 7-8), and excellent (scores ≥9) (43). Secondary outcomes included items evaluating work ability related to physical and mental demands (5-point Likert scale) and estimated future work ability in 2 years (3-point Likert scale), and occupational changes related to PCC during follow-up. We defined PCC using two different measures. First, we defined the presence of PCC as participants (self-)reporting any COVID-19 related symptom out of a list of 23 common PCC-related symptoms at 12 months of follow-up (*PCC status*). Second, we used a combined measure of whether participants had fully recovered and how they assessed their current health status (using the EuroQol visual analog scale (EQ-VAS)) at 12 months (*non-recovery and health impairment status*); non-recovered participants were categorized into mild (EQ-VAS >70), moderate (EQ-VAS 51–70) and severe health impairment (EQ-VAS ≤50) based on population-normative values from previous research (10,44–46). Further measures indicating potential presence of PCC were individual COVID-19 related symptoms, commonly reported PCC-related symptom clusters (fatigue/physical exertion, cardiorespiratory (defined as dyspnea, palpitation, or chest pain), or neurocognitive (defined as concentration, memory, or sleeping problems)), EuroQol 5-dimension 5-level scale (EQ-5D-5L), Fatigue Assessment Scale (FAS), 21-item Depression, Anxiety and Stress Scale (DASS-21), and modified Medical Research Council (mMRC) dyspnea scale. Further information on evaluated work ability outcomes, occupational changes, and PCC-related outcomes is provided in Supplementary Table S1.

### Statistical Analysis

We descriptively compared the reported work ability outcomes in individuals with PCC or reporting non-recovery with associated level of health impairment 12 months after diagnosis. We further descriptively analyzed differences between individuals reporting individual symptoms, symptom clusters, or problems in any of the standardized health assessments (EQ-5D-5L overall and subdomains, FAS, DASS-21, mMRC dyspnea scale) and those without. We then used multivariable regression models to evaluate the association of PCC-related outcomes with work ability outcomes. Model selection included age, sex, baseline health status, hospitalization during acute infection as a priori covariates, with education level, comorbidity count, and history of psychiatric diagnosis added based on improved model fit using the Bayesian Information criterion (BIC; 2-point change considered relevant). For current work ability, we used linear regression (scores 1–10) in primary and ordinal logistic regression (poor, moderate, excellent work ability) in sensitivity analyses. We used ordinal logistic regression for Likert scale-based work ability outcomes. Correspondingly, we report adjusted linear model estimates and odds ratios (ORs) with corresponding 95% confidence intervals (CIs). We evaluated differences in the strength of association (i.e., effect modification) between participant subgroups based on sex (male vs. female), age (40–64 years vs. 18–39 years), comorbidity count (0–1 comorbidity vs. ≥2 comorbidities), and history of psychiatric diagnosis (present vs. absent), descriptively and by using interaction models. Additionally, we descriptively analyzed differences in work ability based on the occurrence of new or worsened psychiatric diagnoses. And last, we described the occupational changes by participants overall and specifically related to PCC. We performed all statistical analyses using R (v4.2.2) (47).

## Results

### Participant Characteristics

Of 1106 Zurich SARS-CoV-2 Cohort participants, 306 were not part of the working-age population, 15 were excluded due to reinfection, and 113 did not provide data at 12 months (Figure 1). Of 672 participants, included in this study, 364 (54.2%) were female, 390 (58.0%) were aged 40-64 years, 79 (11.8%) were asymptomatic, and 9 (1.3%) were hospitalized at initial infection (Table 1). 19 participants (2.8%) reported being unemployed and 4 (0.6%) reported receiving disability insurance benefits at baseline. There were differences in age, sex, severity of acute infection, comorbidities, and history of psychiatric diagnoses between those categorized as having PCC and those without (Supplementary Table S2).

**Figure 1:**
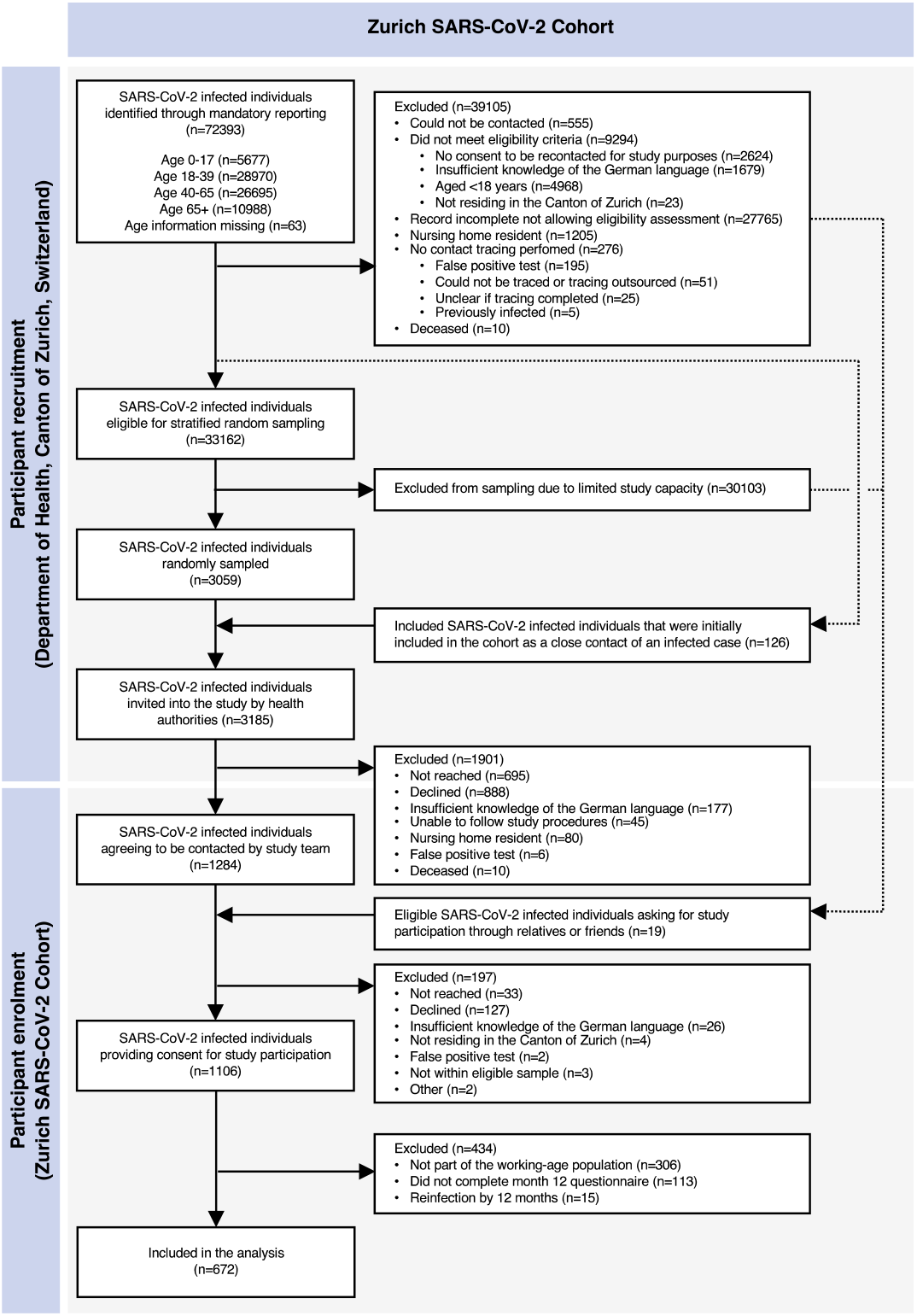
Flow chart of participant enrollment and inclusion in this study.

**Table 1:**
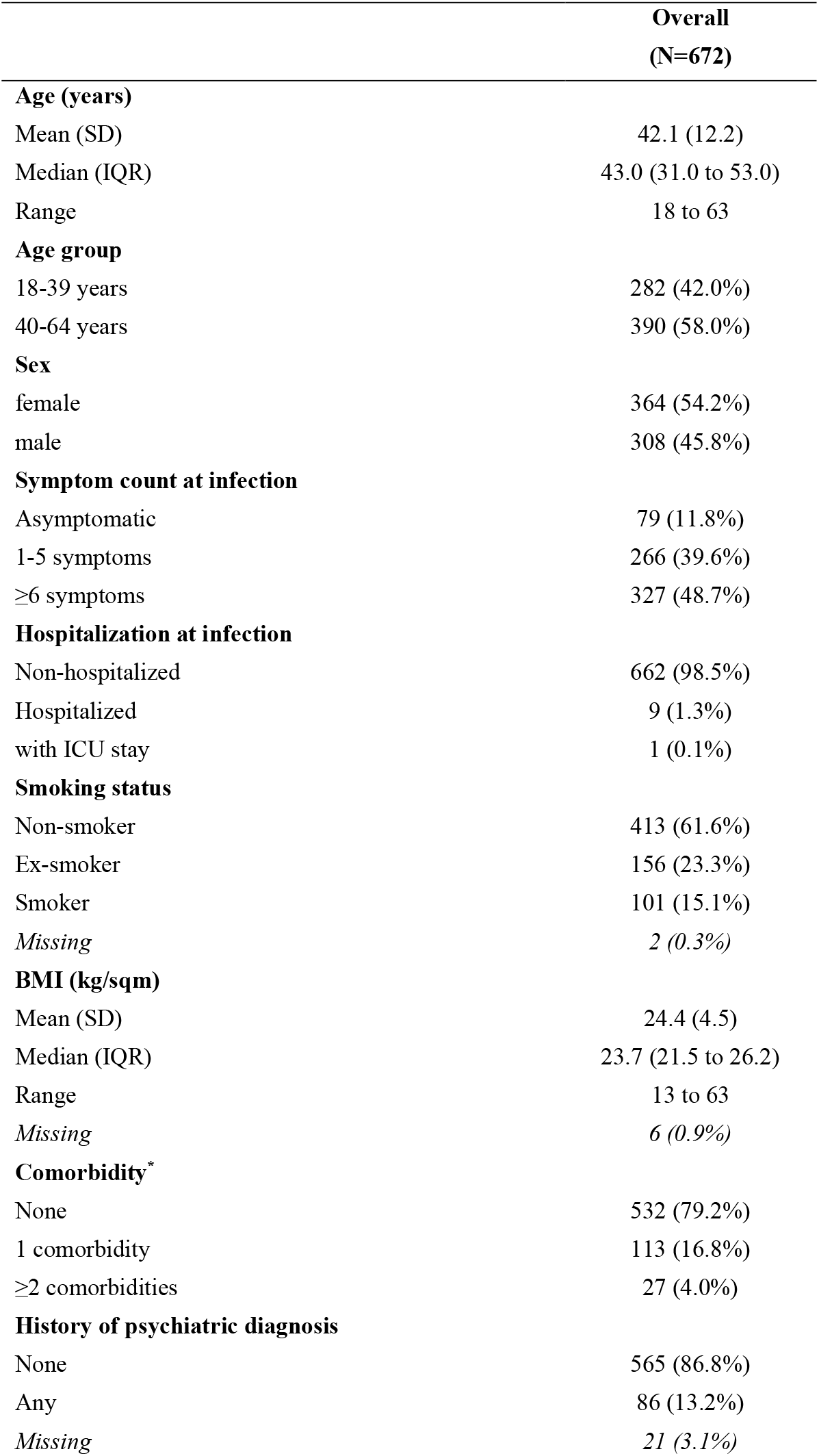
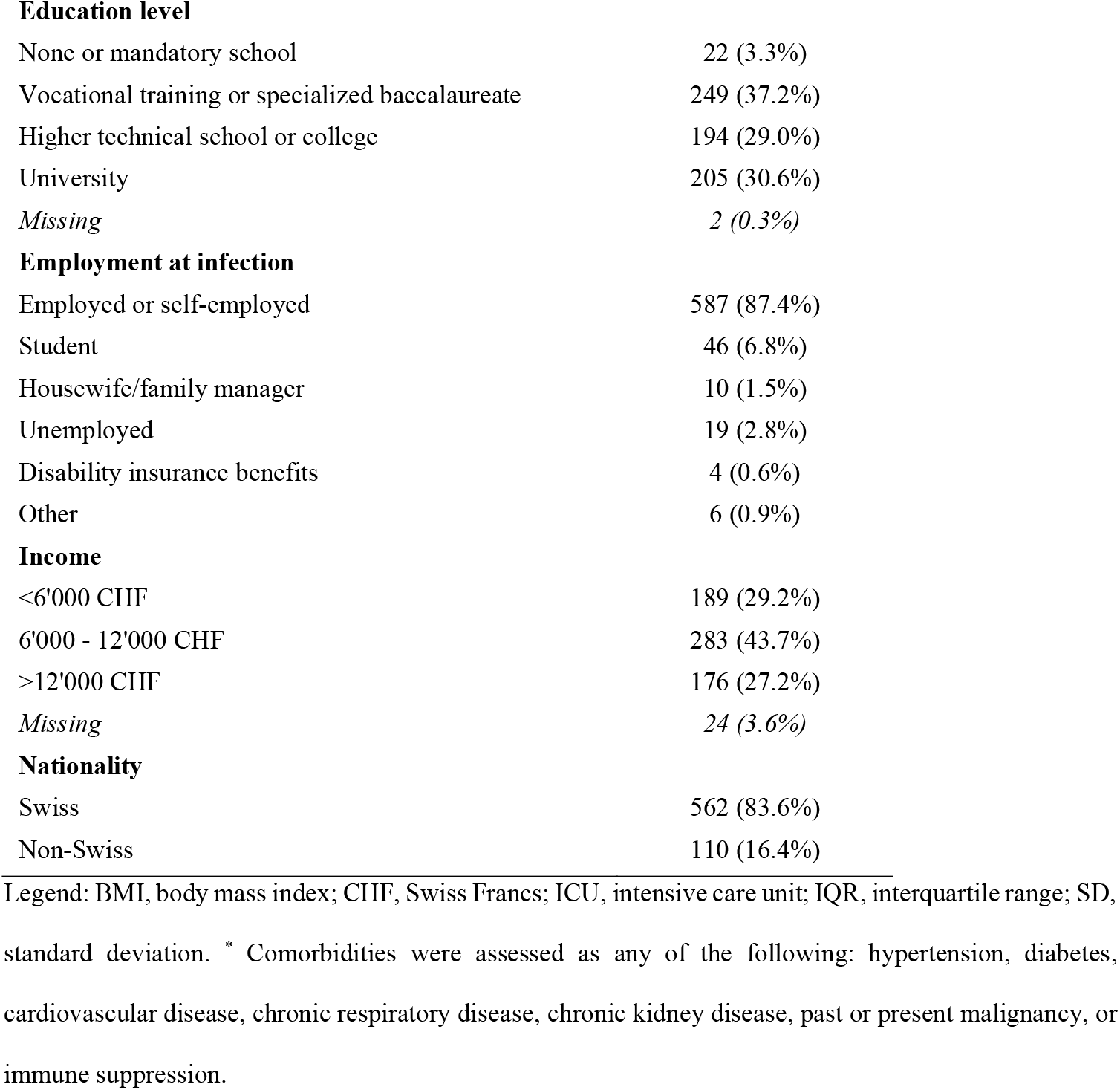
Study participant characteristics.

|  | Overall<br>(N=672) |
| --- | --- |
| **Age (years)** |  |
| Mean (SD) | 42.1 (12.2) |
| Median (IQR) | 43.0 (31.0 to 53.0) |
| Range | 18 to 63 |
| **Age group** |  |
| 18-39 years | 282 (42.0%) |
| 40-64 years | 390 (58.0%) |
| **Sex** |  |
| female | 364 (54.2%) |
| male | 308 (45.8%) |
| **Symptom count at infection** |  |
| Asymptomatic | 79 (11.8%) |
| 1-5 symptoms | 266 (39.6%) |
| ≥6 symptoms | 327 (48.7%) |
| **Hospitalization at infection** |  |
| Non-hospitalized | 662 (98.5%) |
| Hospitalized | 9 (1.3%) |
| with ICU stay | 1 (0.1%) |
| **Smoking status** |  |
| Non-smoker | 413 (61.6%) |
| Ex-smoker | 156 (23.3%) |
| Smoker | 101 (15.1%) |
| Missing | 2 (0.3%) |
| **BMI (kg/sqm)** |  |
| Mean (SD) | 24.4 (4.5) |
| Median (IQR) | 23.7 (21.5 to 26.2) |
| Range | 13 to 63 |
| Missing | 6 (0.9%) |
| **Comorbidity\*** |  |
| None | 532 (79.2%) |
| 1 comorbidity | 113 (16.8%) |
| ≥2 comorbidities | 27 (4.0%) |
| **History of psychiatric diagnosis** |  |
| None | 565 (86.8%) |
| Any | 86 (13.2%) |
| Missing | 21 (3.1%) |
| **Education level** |  |
| None or mandatory school | 22 (3.3%) |
| Vocational training or specialized baccalaureate | 249 (37.2%) |
| Higher technical school or college | 194 (29.0%) |
| University | 205 (30.6%) |
| *Missing* | 2 (0.3%) |
| **Employment at infection** |  |
| Employed or self-employed | 587 (87.4%) |
| Student | 46 (6.8%) |
| Housewife/family manager | 10 (1.5%) |
| Unemployed | 19 (2.8%) |
| Disability insurance benefits | 4 (0.6%) |
| Other | 6 (0.9%) |
| **Income** |  |
| <6'000 CHF | 189 (29.2%) |
| 6'000 - 12'000 CHF | 283 (43.7%) |
| >12'000 CHF | 176 (27.2%) |
| *Missing* | 24 (3.6%) |
| **Nationality** |  |
| Swiss | 562 (83.6%) |
| Non-Swiss | 110 (16.4%) |

Legend: BMI, body mass index; CHF, Swiss Francs; ICU, intensive care unit; IQR, interquartile range; SD, standard deviation. \* Comorbidities were assessed as any of the following: hypertension, diabetes, cardiovascular disease, chronic respiratory disease, chronic kidney disease, past or present malignancy, or immune suppression.

### Work Ability

In descriptive analyses of current work ability, ability related to physical and mental demands at work, and estimated future work ability in 2 years, there was a relevant reduction in work ability across all four outcomes among those with PCC (based on reporting COVID-19 related symptoms) compared to those without and among those reporting non-recovery compared to those that had recovered at 12 months (Figure 2, Supplementary Table S3). Work ability among those reporting non-recovery was more strongly reduced in those with moderate and severe health impairment compared to those with mild health impairment.

**Figure 2:**
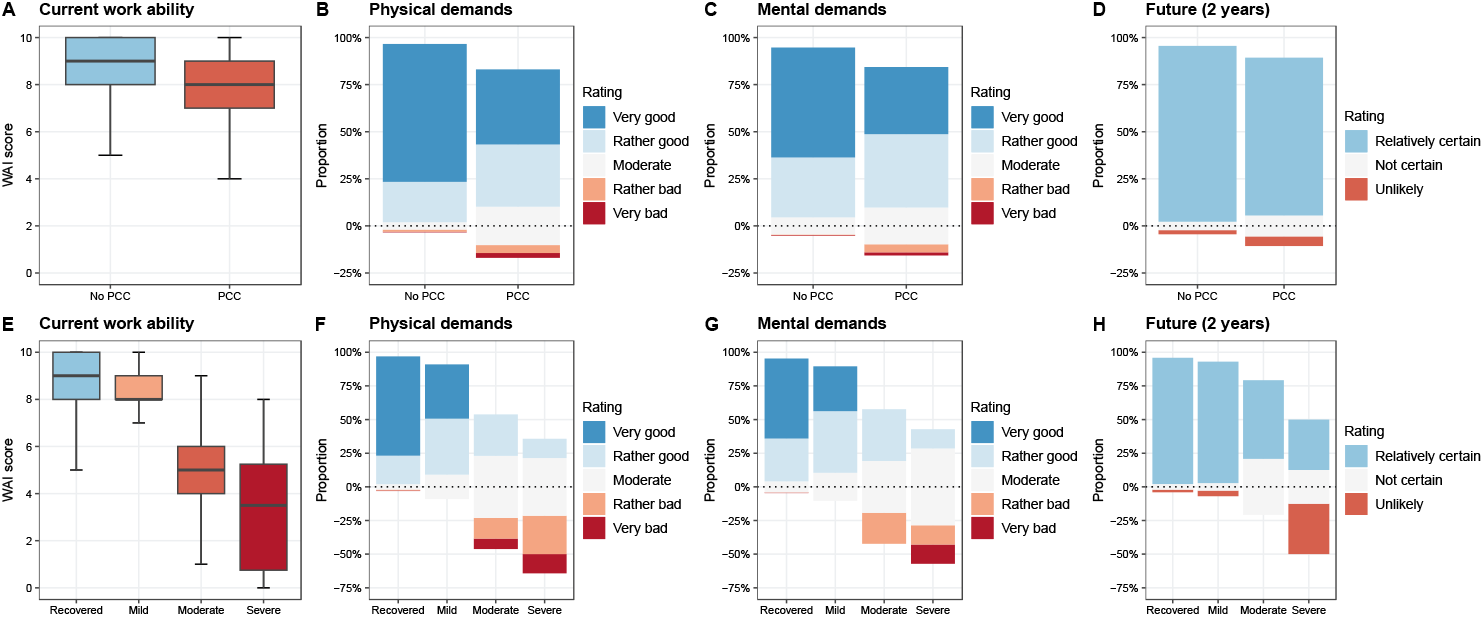
Current work ability, work ability related to physical and mental demands, and estimated future work ability in 2 years by presence of post COVID-19 condition and non-recovery and health impairment at 12 months after diagnosis of primary infection. Panels A–D demonstrate the level of current work ability (A), work ability related to physical (B) and mental (C) demands, and estimated work ability in 2 years (D) between individuals with post COVID-19 condition (PCC)-related symptoms at 12 months compared to those without PCC. Panels E–H show the level of current work ability (E), work ability related to physical (F) and mental (G) demands, and estimated work ability in 2 years (H) between individuals reporting non-recovery with mild, moderate, or severe health impairment at 12 months compared to those reporting full recovery at 12 months. Legend: PCC, post COVID-19 condition; WAI, work ability index.

In adjusted regression analyses, there was very strong evidence that current work ability scores were 0.62 (95% CI 0.30 to 0.95) points lower among those with PCC compared to those without (Figure 3). Current work ability scores were 0.55 (0.21 to 0.88), 3.37 (2.58 to 4.16), and 5.10 (4.16 to 6.04) points lower among those with non-recovery and mild, moderate, and severe health impairment, respectively, compared to those reporting full recovery (very strong evidence). Similarly, there was very strong evidence for a lower odds of having higher work ability with respect to physical (OR 0.30, 95% CI 0.20 to 0.46) and mental (OR 0.40, 0.27 to 0.62) demands among those with PCC compared to those without. Results were similar when evaluating non-recovered individuals compared to those reporting recovery, while reductions in work ability were more pronounced with higher levels of health impairment. There was no evidence for lower odds of having higher estimated future work ability in 2 years (OR 0.52, 0.26 to 1.06) among those with PCC compared to those without and among those with non-recovery and mild health impairment compared to those reporting recovery, but very strong evidence for a reduction in those with moderate or severe health impairment compared to recovered participants. Sensitivity analyses treating current work ability as an ordinal outcome showed similar results (Supplementary Figure S1).

**Figure 3:**
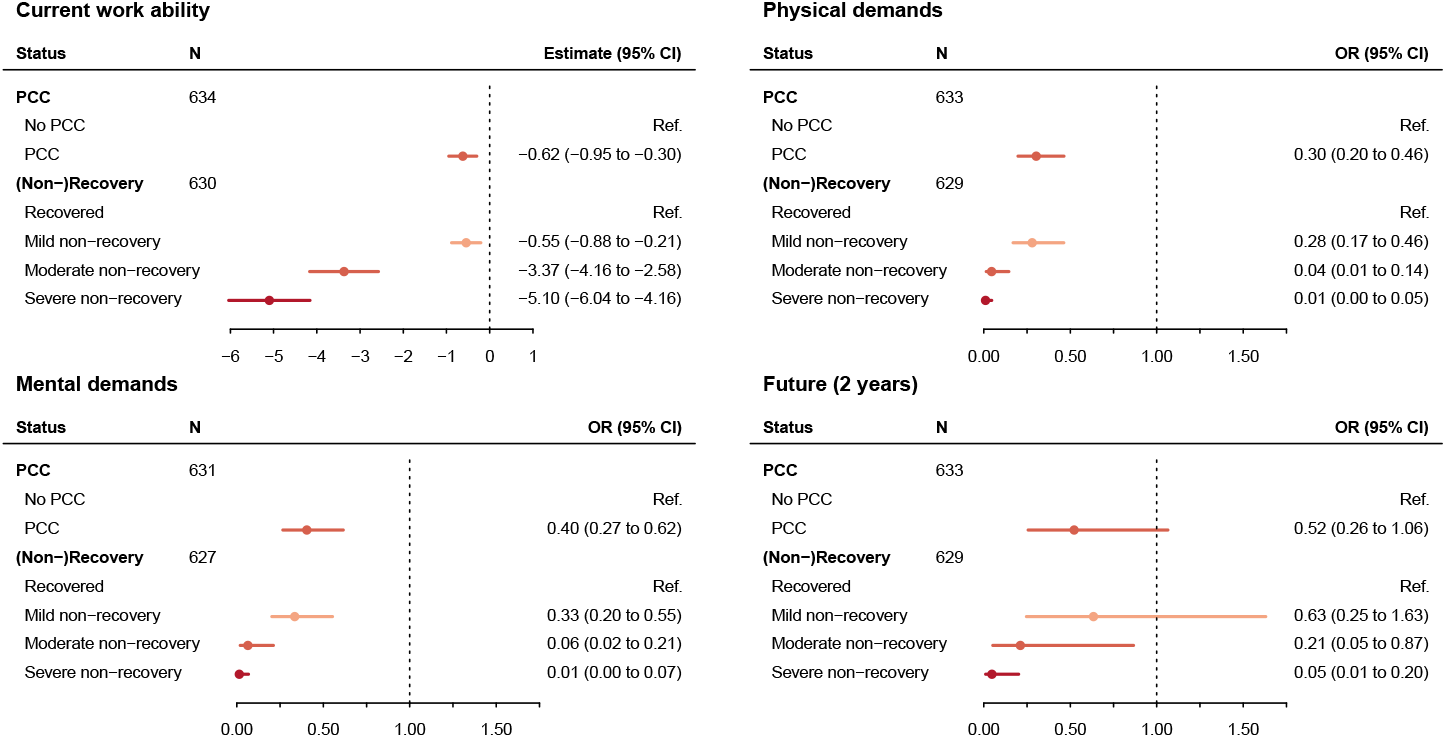
Results from multivariable regression analyses of the association between presence of post COVID-19 condition and current work ability, work ability related to physical and mental demands, and estimated future work ability in 2 years at 12 months after diagnosis of primary infection. Each panel demonstrates results from multivariable linear regression (current work ability) or ordinal logistic regression (work ability related to physical and mental demands, estimated work ability in future) adjusted for sex, age, education level, baseline EuroQol visual analog scale (EQ-VAS), comorbidity count, history of psychiatric diagnosis, and hospitalization at acute infection. Separate models were estimated for the two definitions based on COVID-19 related symptoms (PCC vs. no PCC) and (non-)recovery (severe, moderate or mild health impairment vs. recovery). Legend: CI, confidence interval; OR, odds ratio; PCC, post COVID-19 condition; Ref., reference.

Further analyses demonstrate the association between the presence of specific symptom clusters, individual COVID-19 related symptoms, and presence of health problems in EQ-5D-5L, FAS, DASS-21, and mMRC dyspnea scale and current work ability at 12 months (Supplementary Figures S2–S4, Supplementary Tables S4–S8). Across these analyses, there was very strong evidence for an association between most of the outcomes and current work ability, although not for all individual symptoms.

### Work Ability in Participant Subgroups

In subgroup analyses, there was strong evidence for a difference in the association (i.e., effect modification) of PCC with current work ability and work ability related to physical demands between participants aged 40–64 years and those aged 18–39 years, with a higher reduction in work ability in the older group (Table 2, Supplementary Tables S9–S12). Meanwhile, there was no evidence for a difference in the association of PCC with any work ability outcome between male and female participants, or between participants with 0– 1 comorbidity and participants with ≥2 comorbidities. Last, there was a stronger association of PCC with current work ability (strong evidence) and work ability related to mental demands (weak evidence) in participants with history of psychiatric diagnosis compared to those without. Further descriptive analyses demonstrated relevant differences between participants with different mental health trajectories, indicating a stronger reduction in work ability among participants with history of psychiatric diagnosis and those with a new or worsened psychiatric diagnosis compared to those without history or new or worsened diagnosis, respectively (Supplementary Table S13).

**Table 2:**
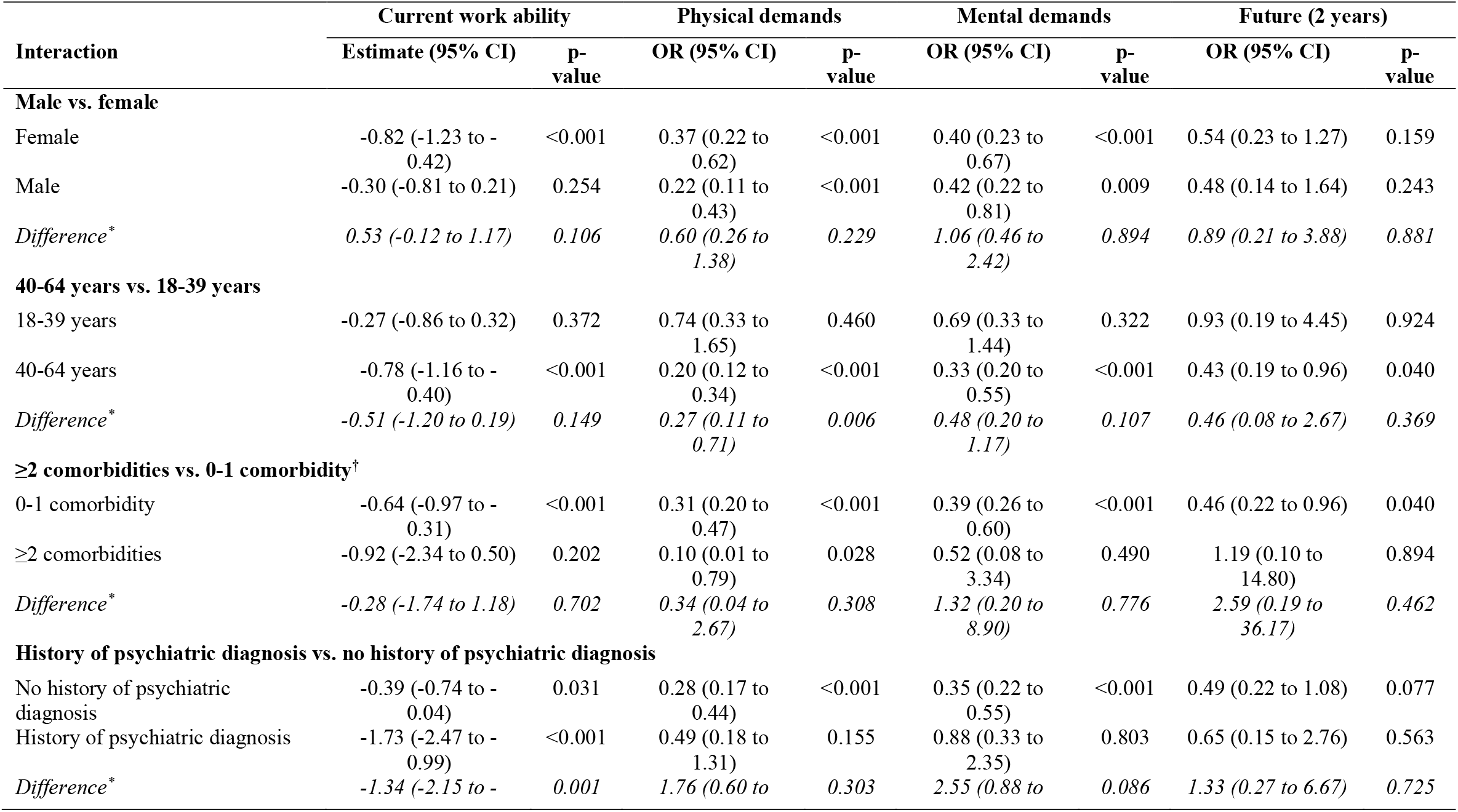
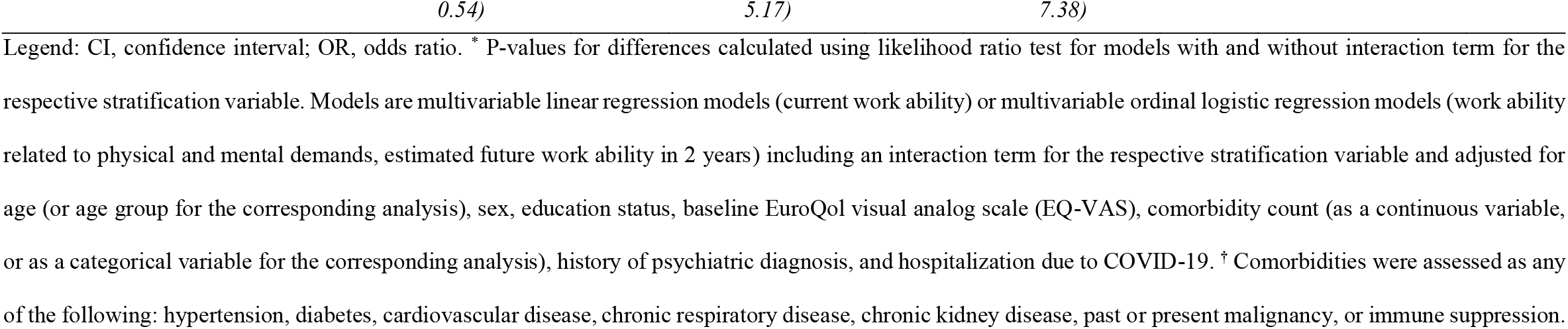
Results from multivariable regression analyses for the association of post COVID-19 condition (PCC, defined as presence of COVID-19 related symptoms) with work ability outcomes (PCC vs. no PCC) within subgroups based on sex, age group, comorbidity count, or history of psychiatric diagnosis.

### Occupational Changes

When evaluating occupational changes up to 12 months, overall 119 (18.1%) participants reported to have had such a change during follow-up (Table 3), with a slightly higher proportion among participants with PCC (31/120, 25.8%) compared to those without PCC (88/552, 16.3%). 7 participants (1.1% of all participants, 5.8% of those with PCC) reported to have faced direct effects by PCC on their occupational situation. Work ability at 12 months was relevantly reduced among those 7 participants with occupational changes related to PCC compared to those without occupational changes and those with PCC-unrelated occupational changes (Supplementary Table S14).

**Table 3:** Occupational changes related to post COVID-19 condition and overall, stratified by post COVID-19 condition and (non-)recovery and health impairment status.

|  | PCC status |  | (Non-)recovery and health impairment status |  |  |  | Overall |
| --- | --- | --- | --- | --- | --- | --- | --- |
|  | No PCC | PCC | Recovered | Mild | Moderate | Severe |  |
|  | (N=552) | (N=120) | (N=562) | (N=72) | (N=13) | (N=8) | (N=672) |
| **Occupational change** |  |  |  |  |  |  |  |
| No occupational change | 451 (83.7%) | 89 (74.2%) | 468 (83.9%) | 58 (80.6%) | 6 (46.2%) | 3 (37.5%) | 540 (81.9%) |
| Occupational change unrelated to PCC | 88 (16.3%) | 24 (20.0%) | 90 (16.1%) | 13 (18.1%) | 4 (30.8%) | 3 (37.5%) | 112 (17.0%) |
| Occupational change related to PCC | 0 (0.0%) | 7 (5.8%) | 0 (0.0%) | 1 (1.4%) | 3 (23.1%) | 2 (25.0%) | 7 (1.1%) |
| *Missing* | *13 (2.4%)* | *0 (0%)* | *4 (0.7%)* | *0 (0%)* | *0 (0%)* | *0 (0%)* | *13 (1.9%)* |
| **Reason for occupational change\*** |  |  |  |  |  |  |  |
| Retired | 4 (4.6%) | 2 (6.5%) | 5 (5.6%) | 0 (0.0%) | 0 (0.0%) | 1 (20.0%) | 6 (5.1%) |
| On permanent sick leave | 3 (3.4%) | 1 (3.2%) | 2 (2.2%) | 0 (0.0%) | 1 (14.3%) | 0 (0.0%) | 4 (3.4%) |
| Receiving disability benefits | 0 (0.0%) | 1 (3.2%) | 0 (0.0%) | 0 (0.0%) | 1 (14.3%) | 0 (0.0%) | 1 (0.8%) |
| Work leave for different reason | 3 (3.4%) | 0 (0.0%) | 3 (3.4%) | 0 (0.0%) | 0 (0.0%) | 0 (0.0%) | 3 (2.5%) |
| Newly self-employed | 2 (2.3%) | 0 (0.0%) | 2 (2.2%) | 0 (0.0%) | 0 (0.0%) | 0 (0.0%) | 2 (1.7%) |
| Changed workplace | 42 (48.3%) | 14 (45.2%) | 46 (51.7%) | 7 (50.0%) | 3 (42.9%) | 0 (0.0%) | 56 (47.5%) |
| Changed position within same workplace | 13 (14.9%) | 4 (12.9%) | 11 (12.4%) | 4 (28.6%) | 0 (0.0%) | 1 (20.0%) | 17 (14.4%) |
| Started training or university studies | 5 (5.7%) | 0 (0.0%) | 4 (4.5%) | 1 (7.1%) | 0 (0.0%) | 0 (0.0%) | 5 (4.2%) |
| Reduced working hours | 2 (2.3%) | 0 (0.0%) | 2 (2.2%) | 0 (0.0%) | 0 (0.0%) | 0 (0.0%) | 2 (1.7%) |
| Lost employment | 4 (4.6%) | 2 (6.5%) | 5 (5.6%) | 0 (0.0%) | 1 (14.3%) | 0 (0.0%) | 6 (5.1%) |
| Other | 9 (10.3%) | 7 (22.6%) | 9 (10.1%) | 2 (14.3%) | 1 (14.3%) | 3 (60.0%) | 16 (13.6%) |
| *Missing* | *1 (1.1%)* | *0 (0%)* | *1 (1.1%)* | *0 (0%)* | *0 (0%)* | *0 (0%)* | *1 (0.8%)* |
| **Financial difficulties due to occupational change\*** |  |  |  |  |  |  |  |
| No | 57 (64.8%) | 17 (54.8%) | 56 (62.2%) | 10 (71.4%) | 4 (57.1%) | 3 (60.0%) | 74 (62.2%) |
| Rather not | 12 (13.6%) | 6 (19.4%) | 15 (16.7%) | 3 (21.4%) | 0 (0.0%) | 0 (0.0%) | 18 (15.1%) |
| Yes, a little | 14 (15.9%) | 7 (22.6%) | 15 (16.7%) | 1 (7.1%) | 3 (42.9%) | 1 (20.0%) | 21 (17.6%) |
| Yes, very much | 5 (5.7%) | 0 (0.0%) | 4 (4.4%) | 0 (0.0%) | 0 (0.0%) | 0 (0.0%) | 5 (4.2%) |
| Unclear/no answer | 0 (0.0%) | 1 (3.2%) | 0 (0.0%) | 0 (0.0%) | 0 (0.0%) | 1 (20.0%) | 1 (0.8%) |

Legend: PCC, post COVID-19 condition. \* Percentages calculated within total of individuals with any occupational change (N=119).

The 7 participants with PCC-related occupational changes reported various individual stories in how PCC affected their work life. One participant lost their work due to PCC. Another reported to be on permanent sick leave at 12 months and being severely affected in daily life. One participant reported that they were unemployed at baseline and could not take on a new position due to PCC, and one was in a job re-integration program but was unable to re-enter the job market due to PCC. Another participant was so severely impacted cognitively that they could no longer use their professional skills (university level) and had to switch to doing simple administrative tasks. One health care worker reported that they had to take a different position that did not require working night shifts. And one participant reported that they had to discontinue their self-employed work as an instructor and seek another part-time job to cope financially because of PCC. Overall, 3 participants (43%) with PCC-related occupational changes reported to have some financial difficulties as a result of their condition and their resulting occupational situation.

## Discussion

### Main Findings in Context

In this prospective population-based cohort of working-age individuals previously infected with SARS-CoV-2, we found that the presence of PCC was strongly associated with a reduction in work ability at 12 months after diagnosis. Among non-recovered, higher levels of health impairment were also associated with substantially lower current work ability and work ability related to physical and mental demands. We found strong evidence that higher age and a history of psychiatric diagnosis was associated with a stronger reduction in current work ability. About 1 in 15 of those with PCC reported having had occupational changes due to PCC within one year, with 1.6% completely dropping out of the workforce.

Evidence on the impact of PCC on the occupational situation and work-related impairments due to PCC is limited and heterogenous (17–27). Prior studies have reported that between 11% and 50% of individuals do not return to work (2,14,26) and that 10% to 72% do not fully regain their work capacity 6 to 12 months after infection (7,23,26,27). Our estimate of 5.8% with occupational changes related to PCC falls in the lower bound of these estimates, which is likely explained by differences in the evaluated populations (e.g., many studies focused on healthcare workers or severely ill patients) and timepoint of assessment (very few with follow-up six months or longer). Differences between countries in terms of sickness and disability benefits systems, as well as cultural and organizational factors, may also explain the wide range of estimates in the literature. Nevertheless, the impact of PCC on the working-age population appears to be substantial and will likely lead to long-term burdens on economic and healthcare systems.

An important factor that determines sustainable return to work is the perceived work ability, which is also more independent of the specific context than return to work and occupational changes. To date, few studies have evaluated work ability in the context of PCC within specific populations of health-care workers and patients attending a post COVID-19 clinic (17,22). Evidence from this study and other studies demonstrated lower work ability scores among those with PCC, with a higher reduction among those with occupational changes. However, it is important to note that although most of the participants with PCC did not have occupational changes and remained at work, decreased work ability in this group may still indicate reduced productivity and efficiency. Sickness presenteeism (i.e., continuing to work while sick) may have negative effects on both the individuals and their employers (48). Sick employees usually need extra efforts to cope with job demands which may lead to additional worsening of their health, and the costs of having a sick employee are estimated to be the same as or even higher than their actual absence (48). Strategies that improve work-related capacity in individuals affected by PCC and promote return to work are urgently needed. In addition, since reduced work ability also is a predictor of early retirement (38,39), it will be vital in the coming years to continuously monitor whether there are increases in the number of people retiring early due to PCC.

In line with other studies, we found a more substantial decrease in current work ability among individuals aged 40-64 years compared to younger individuals. This is concerning since the middle-aged population is typically viewed as the foundation of most economies, as they account for a significant proportion of the workforce, tax revenue, and gross domestic product. We also found that individuals with a history of psychiatric diagnosis had a greater reduction in work ability than those without. The relationship between work and mental health is well-established in the literature (30,49). Targeted strategies and support measures from occupational and rehabilitation medicine, possibly leveraging pre-existing programs for individuals with chronic illnesses, should be put in place to support individuals affected by PCC. In addition, both employees and employers need to be made aware of the mental health aspects of PCC and the impact of mental health on work, as health-promoting working conditions and, for example, supportive leadership may be relevant to the re-integration of relevant subgroups of employees (50).

Fallout from reduced work capacity results not only in financial and health challenges for individuals affected by PCC, but can also have substantial consequences for the economy and society in the longer term. Altogether, our findings underline the necessity for interdisciplinary interventions aimed at individuals affected by PCC, including those with moderate or even mild health impairment. Given that early intervention is a core principle of occupational rehabilitation, further research is warranted to determine whether earlier rehabilitation could improve work outcomes in people with persistent symptoms after COVID-19 but who are not yet diagnosed with PCC. Identifying specific COVID-19 symptoms that predict impairment in work ability will help to develop and provide such early interventions. We consider this study to be part of that effort.

### Limitations

Strengths of the study include its population-based approach, the large sample size, and the high retention rate at one year (90%) limiting emigrative selection bias arising from loss to follow-up. In addition, the granularity of the data and the use of a validated, internationally used, and context-independent measure of work ability strengthens our evaluation. However, some limitations need to be considered. First, immigrative selection may have occurred if individuals who were more health literate were more likely to participate or if individuals who had PCC and more severely impacted were more likely to be retained in the study. This may have led to an overestimation of the impact of PCC on work ability. In contrast, our findings may be biased towards lower estimates since only a small proportion of the participants were hospitalized for COVID-19. Second, the relatively low proportion of hospitalized participants also limits the generalizability of our results to those with the most severe acute disease, who may also suffer from more severe medical complications and sequelae of the hospital stay (e.g., post intensive care syndrome). Additionally, the generalizability of our findings to individuals infected with emerging SARS-CoV-2 variants of concern or who were vaccinated prior to infection is limited, since our participants were all infected with wildtype SARS-CoV-2 and unvaccinated at infection. The risk of PCC and severe health impairment is substantially reduced with vaccination and infection with newer variants, but still present (51–54). As the impact of PCC on work ability is likely comparable in these contexts, this may have significant socioeconomic implications given that more than 45% of the global population is estimated to have been infected with the Omicron variant (55). Further research is needed to evaluate whether similar reduced work ability and occupational changes are observed in vaccinated populations and in the context of emerging variants of concern. Nonetheless, the population from the early stages of the pandemic included in this study remains highly relevant since these are the individuals experiencing long-term health consequences at present, posing a challenge to public health. Third, we assessed PCC using self-reported measures. Since we could not conduct a clinical validation of PCC, we cannot fully exclude that reported symptoms and health impairment were related to the presence or worsening of other infections or conditions. Meanwhile, self-reported measures are key for capturing the lived experience of those affected, and the comparable results across two different definitions of PCC strengthen the credibility of our findings. Fourth, we did not have data on participants’ work ability prior to SARS-CoV-2 infection. Thus, we cannot be fully certain that the reduced work ability is entirely due to infection and not other preexisting conditions. However, we at least partially accounted for this in our models by adjusting for baseline health status, and the detailed evaluation of the individual stories supports our finding of a reduced work ability related to PCC.

## Conclusion

In this population-based study, we found that PCC significantly reduced the work ability of a relevant proportion of individuals a year after SARS-CoV-2 infection and in some instances led to an inability to work altogether. Such loss of productivity and incapacity to work can have severe implications for individuals, families, and society as a whole. It is critical that policymakers, healthcare professionals, and employers recognize the impact of PCC on the workforce and develop effective strategies and interventions that can support and enable affected individuals in regaining and retaining their work ability.

## Data Availability

All data produced in the present study are available upon reasonable request to the corresponding author after the peer-reviewed publication of the article.

## Declarations

### Ethical Approval

The study was approved by the responsible ethics committee of the Canton of Zurich, Switzerland (BASEC-Nr. 2020-01739). Written or electronic informed consent was obtained from all participants.

### Author Contributions

TB, DM, JSF, and MAP conceived and planned the Zurich SARS-CoV-2 Cohort study. TB, DM, and MAP coordinated the Zurich SARS-CoV-2 Cohort study. PK, TB, MAP, and DM conceived and designed this analysis. TB, DM, AD, and MAP contributed to participant recruitment and data collection. MAP supervised the project. JSF and MAP obtained funding. TB and DM prepared the analytic datasets. DM performed the statistical analysis and SH provided input on the statistical analysis. All authors contributed to the interpretation of the data. PK, TB, and DM wrote the draft manuscript. All authors critically revised and provided feedback on the draft manuscript. All authors read and approved the final manuscript.

## Acknowledgements

We thank the study administration team and the study participants for their continued and highly valuable support.

