## Supplementary Material for "Post COVID-19 Condition, Work Ability and Occupational Changes: Results from a Population-based Cohort"

### Supplementary Methods

**Supplementary Table S1. Details on outcome measurement and definitions.**

| Outcome | Instrument & question wording | Definition/Categories |
|---|---|---|
| **Work ability and occupational changes** | | |
| Current work ability | Work ability index (1–3)<br>• *"Assume that your work ability at its best has a value of 10 points. How many points would you give your current work ability? (0 means that you currently cannot work at all) – (10 work ability at its best)"* | Self-perceived current work ability compared to highest work ability ever.<br><br>Categorical classification based on von Bonsdorff et al.(4):<br>• Poor (score ≤6)<br>• Moderate (score 7-8)<br>• Excellent (score ≥9) |
| Work ability related to physical demands | Work ability index (1–3)<br>• *"How do you rate your current work ability with respect to the physical demands of your work? (Likert scale from very good to very poor)"* | Self-perceived current work ability in relation to physical demands of work. |
| Work ability related to mental demands | Work ability index (1–3)<br>• *"How do you rate your current work ability with respect to the mental demands of your work? (Likert scale from very good to very poor)"* | Self-perceived current work ability in relation to mental demands of work. |
| Estimated future work ability in 2 years | Work ability index (1–3)<br>• *"Do you believe, according to your present state of health, that you will be able to do your current job two years from now? (Unlikely, not certain, relatively certain)"* | Self-perceived estimated work ability in 2 years based on current health status. |
| Occupational changes | • *"Has your occupational situation changed in the last six months? (Yes, No)"*<br>• *"How has your professional situation changed? (I have retired, I am on permanent sick leave, I am on work leave for a different reason, I have started self-employment, I work at a new workplace, I work in a new position within the same* | Question regarding changes in occupational situation was asked at 6 months and 12 months after infection.<br><br>Occupational changes related to post COVID-19 condition were determined based on a pre-defined list of changes and further information provided through comments in free text fields. Phone interviews were conducted for all |

| Outcome | Instrument & question wording | Definition/Categories |
|---|---|---|
| | *workplace, I have started professional training or university studies, I am temporarily out of work due to lack of demand, I have lost my employment, Other reason (specify))"* | participants with PCC where no further information on the specific occupational change was provided by the participant or where the relation to COVID-19 was unequivocal. |
| **Post COVID-19 condition** | | |
| PCC status | • *"In the past 7 days, have you had one or more of the following symptoms that are unrelated to a chronic illness or allergy? (Fatigue, Post-exertional malaise, Fever, Cough, Dyspnea or shortness of breath, Chest pain, Heart palpitations, Altered taste and/or smell, Headache, Concentration difficulties, Memory problems, Vertigo or dizziness, Tremors, Tingling sensation of extremities, Sleep disturbances, Myalgia Arthralgia, Gastrointestinal disturbances, Swallowing difficulties, Hearing problems, Visual disturbances, Skin rash, Hair loss)"*<br>• *"Do you think this symptom is related to or a complication of coronavirus disease "COVID-19"? (No, Yes, I don't know)"* | Presence of at least one symptom in a list of 23 common PCC-related symptoms that were reported by participants to be related to PCC. |
| Non-recovery and health impairment status | • Self-reported recovery status: *"How do you feel now compared to when you were infected? (fully recovered and symptom free, better but not fully recovered, neither better nor worse, worse)"*<br>• EQ-VAS | 1. **Recovered**: participants reporting to fully recovered to their normal health status and symptom-free, or unchanged if asymptomatic at infection.<br>2. **Non-recovered with mild health impairment**: participants reporting not to be back to their normal health status and reporting EQ-VAS scores >70.<br>3. **Non-recovered with moderate health impairment**: participants reporting not to be back to their normal health status and |

| Outcome | Instrument & question wording | Definition/Categories |
|---|---|---|
| | | reporting EQ-VAS scores between 51–70.<br>4. **Non-recovered with severe health impairment**: participants reporting not to be back to their normal health status and reporting EQ-VAS scores ≤50.<br><br>EQ-VAS cut-offs were determined based on population-normative values from prior research (5–8). |
| Individual PCC-related symptoms | • *«In the past 7 days, have you had one or more of the following symptoms that are unrelated to a chronic illness or allergy? (symptom list as above)"*<br>• *"Do you think this symptom is related to or a complication of coronavirus disease "COVID-19"? (No, Yes, I don't know)"* | Presence of each individual symptom in a list of 23 common PCC-related symptoms (see above) that were reported by participants to be related to PCC. |
| Symptom clusters | • Fatigue/physical exertion (defined as presence of fatigue or physical exertion)<br>• Cardiorespiratory (defined as presence of dyspnea, palpitation, or chest pain)<br>• Neurocognitive (defined as concentration, memory, or sleeping problems) | Presence of symptoms belonging to each symptom cluster. |
| Health-related quality of life (HRQoL) | EQ-5D-5L | Presence of problems in 5 dimensions (mobility, self-care, usual activities, depression or anxiety, pain or discomfort). |
| Fatigue | FAS | Presence of fatigue (defined as FAS score ≥22). |
| Depression, anxiety or stress | DASS-21 | Presence of depression (defined as score ≥10 in depression-related items), anxiety (score ≥8 in anxiety-related items), or stress (score ≥15 in stress-related items). |
| Dyspnea | mMRC dyspnea scale | Degree of functional disability due to dyspnea (defined as mMRC dyspnea grade ≥1). |

Legend: DASS-21, 21-item Depression, Anxiety and Stress Scale; EQ-VAS, EuroQol visual analog scale; EQ-5D-5L, EuroQol 5-dimension 5-level scale; FAS, Fatigue Assessment Scale; mMRC, modified Medical Research Council scale; PCC, post COVID-19 condition.

### Supplementary Results

**Supplementary Table S2: Detailed study population characteristics, stratified by presence of COVID-19 related symptoms (post COVID-19 condition) and (non-)recovery and health impairment status at 12 months.**

| | PCC status | | (Non-)recovery and health impairment status | | | | Overall |
|---|---|---|---|---|---|---|---|
| | No PCC (N=552) | PCC (N=120) | Recovered (N=562) | Mild (N=72) | Moderate (N=13) | Severe (N=8) | (N=672) |
| **Age (years)** | | | | | | | |
| Mean (SD) | 41.1 (12.2) | 46.6 (10.9) | 41.2 (12.2) | 46.0 (11.4) | 49.2 (8.8) | 49.5 (12.8) | 42.1 (12.2) |
| Median (IQR) | 41.0 (30.8 to 51.2) | 50.0 (38.0 to 55.0) | 41.5 (30.0 to 51.8) | 48.0 (35.8 to 55.2) | 51.0 (45.0 to 56.0) | 53.0 (45.0 to 58.0) | 43.0 (31.0 to 53.0) |
| Range | 18 to 63 | 21 to 62 | 18 to 63 | 25 to 62 | 32 to 59 | 24 to 62 | 18 to 63 |
| **Age group** | | | | | | | |
| 18-39 years | 249 (45.1%) | 33 (27.5%) | 250 (44.5%) | 22 (30.6%) | 3 (23.1%) | 2 (25.0%) | 282 (42.0%) |
| 40-64 years | 303 (54.9%) | 87 (72.5%) | 312 (55.5%) | 50 (69.4%) | 10 (76.9%) | 6 (75.0%) | 390 (58.0%) |
| **Sex** | | | | | | | |
| female | 285 (51.6%) | 79 (65.8%) | 292 (52.0%) | 47 (65.3%) | 11 (84.6%) | 6 (75.0%) | 364 (54.2%) |
| male | 267 (48.4%) | 41 (34.2%) | 270 (48.0%) | 25 (34.7%) | 2 (15.4%) | 2 (25.0%) | 308 (45.8%) |
| **Symptom count at infection** | | | | | | | |
| Asymptomatic | 67 (12.1%) | 12 (10.0%) | 70 (12.5%) | 5 (6.9%) | 1 (7.7%) | 1 (12.5%) | 79 (11.8%) |
| 1-5 symptoms | 233 (42.2%) | 33 (27.5%) | 230 (40.9%) | 25 (34.7%) | 1 (7.7%) | 3 (37.5%) | 266 (39.6%) |
| ≥6 symptoms | 252 (45.7%) | 75 (62.5%) | 262 (46.6%) | 42 (58.3%) | 11 (84.6%) | 4 (50.0%) | 327 (48.7%) |
| **Hospitalization at infection** | | | | | | | |
| Non-hospitalized | 547 (99.1%) | 115 (95.8%) | 559 (99.5%) | 69 (95.8%) | 11 (84.6%) | 7 (87.5%) | 662 (98.5%) |
| Hospitalized | 5 (0.9%) | 5 (4.2%) | 3 (0.5%) | 3 (4.2%) | 2 (15.4%) | 1 (12.5%) | 10 (1.5%) |
| with ICU stay | 0 (0.0%) | 1 (0.8%) | 0 (0.0%) | 0 (0.0%) | 1 (7.7%) | 0 (0.0%) | 1 (0.1%) |

| | | | | | | | |
|---|---|---|---|---|---|---|---|
| **Smoking status** | | | | | | | |
| Non-smoker | 343 (62.4%) | 70 (58.3%) | 347 (62.0%) | 42 (58.3%) | 9 (69.2%) | 6 (75.0%) | 413 (61.6%) |
| Ex-smoker | 123 (22.4%) | 33 (27.5%) | 127 (22.7%) | 20 (27.8%) | 2 (15.4%) | 1 (12.5%) | 156 (23.3%) |
| Smoker | 84 (15.3%) | 17 (14.2%) | 86 (15.4%) | 10 (13.9%) | 2 (15.4%) | 1 (12.5%) | 101 (15.1%) |
| *Missing* | *2 (0.4%)* | *0 (0%)* | *2 (0.4%)* | *0 (0%)* | *0 (0%)* | *0 (0%)* | *2 (0.3%)* |
| **BMI (kg/sqm)** | | | | | | | |
| Mean (SD) | 24.2 (4.3) | 25.6 (5.1) | 24.3 (4.3) | 24.9 (4.7) | 29.9 (7.5) | 22.4 (1.9) | 24.4 (4.5) |
| Median (IQR) | 23.6 (21.5 to 25.9) | 24.8 (22.1 to 28.6) | 23.6 (21.5 to 26.0) | 24.5 (21.7 to 27.0) | 30.4 (26.0 to 31.1) | 22.3 (20.7 to 23.7) | 23.7 (21.5 to 26.2) |
| Range | 13 to 63 | 17 to 45 | 17 to 63 | 18 to 40 | 20 to 45 | 20 to 25 | 13 to 63 |
| *Missing* | *5 (0.9%)* | *1 (0.8%)* | *6 (1.1%)* | *0 (0%)* | *0 (0%)* | *0 (0%)* | *6 (0.9%)* |
| **Comorbidity*** | | | | | | | |
| None | 453 (82.1%) | 79 (65.8%) | 460 (81.9%) | 49 (68.1%) | 6 (46.2%) | 4 (50.0%) | 532 (79.2%) |
| 1 comorbidity | 80 (14.5%) | 33 (27.5%) | 85 (15.1%) | 18 (25.0%) | 5 (38.5%) | 3 (37.5%) | 113 (16.8%) |
| ≥2 comorbidities | 19 (3.4%) | 8 (6.7%) | 17 (3.0%) | 5 (6.9%) | 2 (15.4%) | 1 (12.5%) | 27 (4.0%) |
| **Comorbidity count*** | | | | | | | |
| Median (IQR) | 0 (0 to 0) | 0 (0 to 1) | 0 (0 to 0) | 0 (0 to 1) | 1 (0 to 1) | 0.5 (0 to 1) | 0 (0 to 0) |
| Range | 0 to 3 | 0 to 2 | 0 to 3 | 0 to 2 | 0 to 2 | 0 to 2 | 0 to 3 |
| **History of psychiatric diagnosis** | | | | | | | |
| None | 472 (88.6%) | 93 (78.8%) | 486 (87.7%) | 63 (87.5%) | 6 (50.0%) | 5 (62.5%) | 565 (86.8%) |
| Any | 61 (11.4%) | 25 (21.2%) | 68 (12.3%) | 9 (12.5%) | 6 (50.0%) | 3 (37.5%) | 86 (13.2%) |
| Depression | 27 (4.9%) | 18 (15.0%) | 32 (5.7%) | 5 (6.9%) | 5 (38.5%) | 3 (37.5%) | 45 (6.7%) |
| Anxiety | 12 (2.2%) | 2 (1.7%) | 13 (2.3%) | 0 (0.0%) | 0 (0.0%) | 1 (12.5%) | 14 (2.1%) |
| Burnout | 6 (1.1%) | 3 (2.5%) | 6 (1.1%) | 1 (1.4%) | 2 (15.4%) | 0 (0.0%) | 9 (1.3%) |
| Bipolar disorder | 2 (0.4%) | 0 (0.0%) | 2 (0.4%) | 0 (0.0%) | 0 (0.0%) | 0 (0.0%) | 2 (0.3%) |
| ADHD | 2 (0.4%) | 1 (0.8%) | 2 (0.4%) | 1 (1.4%) | 0 (0.0%) | 0 (0.0%) | 3 (0.4%) |
| PTSD | 4 (0.7%) | 1 (0.8%) | 4 (0.7%) | 0 (0.0%) | 0 (0.0%) | 1 (12.5%) | 5 (0.7%) |

| | | | | | | | |
|---|---|---|---|---|---|---|---|
| Eating disorder | 1 (0.2%) | 1 (0.8%) | 2 (0.4%) | 0 (0.0%) | 0 (0.0%) | 0 (0.0%) | 2 (0.3%) |
| Sleep disorder | 2 (0.4%) | 0 (0.0%) | 2 (0.4%) | 0 (0.0%) | 0 (0.0%) | 0 (0.0%) | 2 (0.3%) |
| Other | 2 (0.4%) | 0 (0.0%) | 2 (0.4%) | 0 (0.0%) | 0 (0.0%) | 0 (0.0%) | 2 (0.3%) |
| *Missing* | *19 (3.4%)* | *2 (1.7%)* | *8 (1.4%)* | *0 (0%)* | *1 (7.7%)* | *0 (0%)* | *21 (3.1%)* |
| **Education level** | | | | | | | |
| None or mandatory school | 17 (3.1%) | 5 (4.2%) | 16 (2.9%) | 4 (5.6%) | 0 (0.0%) | 0 (0.0%) | 22 (3.3%) |
| Vocational training or specialized baccalaureate | 194 (35.1%) | 55 (46.6%) | 199 (35.5%) | 33 (46.5%) | 7 (53.8%) | 3 (37.5%) | 249 (37.2%) |
| Higher technical school or college | 165 (29.9%) | 29 (24.6%) | 173 (30.8%) | 16 (22.5%) | 2 (15.4%) | 2 (25.0%) | 194 (29.0%) |
| University | 176 (31.9%) | 29 (24.6%) | 173 (30.8%) | 18 (25.4%) | 4 (30.8%) | 3 (37.5%) | 205 (30.6%) |
| *Missing* | *0 (0%)* | *2 (1.7%)* | *1 (0.2%)* | *1 (1.4%)* | *0 (0%)* | *0 (0%)* | *2 (0.3%)* |
| **Employment at infection** | | | | | | | |
| Employed or self-employed | 488 (88.4%) | 99 (82.5%) | 494 (87.9%) | 61 (84.7%) | 11 (84.6%) | 4 (50.0%) | 587 (87.4%) |
| Student | 42 (7.6%) | 4 (3.3%) | 45 (8.0%) | 1 (1.4%) | 0 (0.0%) | 0 (0.0%) | 46 (6.8%) |
| Housewife/family manager | 9 (1.6%) | 1 (0.8%) | 9 (1.6%) | 1 (1.4%) | 0 (0.0%) | 0 (0.0%) | 10 (1.5%) |
| Unemployed | 9 (1.6%) | 10 (8.3%) | 10 (1.8%) | 7 (9.7%) | 1 (7.7%) | 1 (12.5%) | 19 (2.8%) |
| Disability insurance benefits | 0 (0.0%) | 4 (3.3%) | 0 (0.0%) | 0 (0.0%) | 1 (7.7%) | 3 (37.5%) | 4 (0.6%) |
| Other | 4 (0.7%) | 2 (1.7%) | 4 (0.7%) | 2 (2.8%) | 0 (0.0%) | 0 (0.0%) | 6 (0.9%) |
| **Income** | | | | | | | |
| <6'000 CHF | 149 (28.0%) | 40 (34.8%) | 156 (28.7%) | 23 (32.9%) | 3 (25.0%) | 4 (50.0%) | 189 (29.2%) |
| 6'000 - 12'000 CHF | 231 (43.3%) | 52 (45.2%) | 234 (43.1%) | 33 (47.1%) | 7 (58.3%) | 1 (12.5%) | 283 (43.7%) |
| >12'000 CHF | 153 (28.7%) | 23 (20.0%) | 153 (28.2%) | 14 (20.0%) | 2 (16.7%) | 3 (37.5%) | 176 (27.2%) |
| *Missing* | *19 (3.4%)* | *5 (4.2%)* | *19 (3.4%)* | *2 (2.8%)* | *1 (7.7%)* | *0 (0%)* | *24 (3.6%)* |
| **Nationality** | | | | | | | |
| Swiss | 465 (84.2%) | 97 (80.8%) | 479 (85.2%) | 56 (77.8%) | 9 (69.2%) | 7 (87.5%) | 562 (83.6%) |
| Non-Swiss | 87 (15.8%) | 23 (19.2%) | 83 (14.8%) | 16 (22.2%) | 4 (30.8%) | 1 (12.5%) | 110 (16.4%) |

Legend: ADHD, attention deficit hyperactivity disorder; BMI, body mass index; CHF, Swiss Francs; ICU, intensive care unit; IQR, interquartile range; PCC, post COVID-19 condition; PTSD, post-traumatic stress disorder; SD, standard deviation. [*] Comorbidities were assessed as any of the following: hypertension, diabetes, cardiovascular disease, chronic respiratory disease, chronic kidney disease, past or present malignancy, or immune suppression.

**Supplementary Table S3: Work ability outcomes, stratified by post COVID-19 condition (defined as presence of COVID-19 related symptoms) and (non-)recovery and health impairment status at 12 months.**

| | PCC status | | (Non-)recovery and health impairment status | | | | Overall |
|---|---|---|---|---|---|---|---|
| | No PCC (N=552) | PCC (N=120) | Recovered (N=562) | Mild (N=72) | Moderate (N=13) | Severe (N=8) | (N=672) |
| **Current work ability (score)** | | | | | | | |
| Mean (SD) | 8.9 (1.5) | 7.9 (2.2) | 8.9 (1.4) | 8.4 (1.0) | 4.9 (2.1) | 3.4 (2.9) | 8.7 (1.7) |
| Median (IQR) | 9.0 (8.0 to 10.0) | 8.0 (7.0 to 9.0) | 9.0 (8.0 to 10.0) | 8.0 (8.0 to 9.0) | 5.0 (4.0 to 6.0) | 3.5 (0.8 to 5.2) | 9.0 (8.0 to 10.0) |
| Range | 0 to 10 | 0 to 10 | 0 to 10 | 5 to 10 | 1 to 9 | 0 to 8 | 0 to 10 |
| *Missing* | *15 (2.7%)* | *1 (0.8%)* | *7 (1.2%)* | *0 (0%)* | *0 (0%)* | *0 (0%)* | *16 (2.4%)* |
| **Current work ability (categorical)** | | | | | | | |
| Poor (score ≤6) | 27 (5.0%) | 20 (16.8%) | 23 (4.1%) | 2 (2.8%) | 10 (76.9%) | 7 (87.5%) | 47 (7.2%) |
| Moderate (score 7-8) | 137 (25.5%) | 42 (35.3%) | 137 (24.7%) | 38 (52.8%) | 2 (15.4%) | 1 (12.5%) | 179 (27.3%) |
| Excellent (score ≥9) | 373 (69.5%) | 57 (47.9%) | 395 (71.2%) | 32 (44.4%) | 1 (7.7%) | 0 (0.0%) | 430 (65.5%) |
| *Missing* | *15 (2.7%)* | *1 (0.8%)* | *7 (1.2%)* | *0 (0%)* | *0 (0%)* | *0 (0%)* | *16 (2.4%)* |
| **Work ability related to physical demands** | | | | | | | |
| Very bad | 2 (0.4%) | 3 (2.5%) | 2 (0.4%) | 0 (0.0%) | 1 (7.7%) | 1 (14.3%) | 5 (0.8%) |
| Rather bad | 6 (1.1%) | 5 (4.2%) | 4 (0.7%) | 0 (0.0%) | 2 (15.4%) | 2 (28.6%) | 11 (1.7%) |
| Moderate | 21 (3.9%) | 24 (20.3%) | 22 (4.0%) | 13 (18.1%) | 6 (46.2%) | 3 (42.9%) | 45 (6.9%) |
| Rather good | 115 (21.4%) | 39 (33.1%) | 118 (21.3%) | 30 (41.7%) | 4 (30.8%) | 1 (14.3%) | 154 (23.5%) |
| Very good | 393 (73.2%) | 47 (39.8%) | 409 (73.7%) | 29 (40.3%) | 0 (0.0%) | 0 (0.0%) | 440 (67.2%) |
| *Missing* | *15 (2.7%)* | *2 (1.7%)* | *7 (1.2%)* | *0 (0%)* | *0 (0%)* | *1 (12.5%)* | *17 (2.5%)* |
| **Work ability related to mental demands** | | | | | | | |
| Very bad | 2 (0.4%) | 2 (1.7%) | 2 (0.4%) | 0 (0.0%) | 0 (0.0%) | 1 (14.3%) | 4 (0.6%) |

| | | | | | | | |
|---|---|---|---|---|---|---|---|
| Rather bad | 2 (0.4%) | 5 (4.2%) | 1 (0.2%) | 0 (0.0%) | 3 (23.1%) | 1 (14.3%) | 7 (1.1%) |
| Moderate | 49 (9.2%) | 23 (19.5%) | 45 (8.1%) | 15 (20.8%) | 5 (38.5%) | 4 (57.1%) | 72 (11.0%) |
| Rather good | 170 (31.8%) | 46 (39.0%) | 176 (31.8%) | 33 (45.8%) | 5 (38.5%) | 1 (14.3%) | 216 (33.1%) |
| Very good | 312 (58.3%) | 42 (35.6%) | 329 (59.5%) | 24 (33.3%) | 0 (0.0%) | 0 (0.0%) | 354 (54.2%) |
| *Missing* | *17 (3.1%)* | *2 (1.7%)* | *9 (1.6%)* | *0 (0%)* | *0 (0%)* | *1 (12.5%)* | *19 (2.8%)* |
| **Estimated work ability in future (2 years)** | | | | | | | |
| Unlikely | 12 (2.2%) | 6 (5.1%) | 11 (2.0%) | 3 (4.2%) | 0 (0.0%) | 3 (37.5%) | 18 (2.8%) |
| Not certain | 24 (4.5%) | 13 (11.1%) | 23 (4.2%) | 4 (5.6%) | 5 (41.7%) | 2 (25.0%) | 37 (5.7%) |
| Relatively certain | 501 (93.3%) | 98 (83.8%) | 520 (93.9%) | 65 (90.3%) | 7 (58.3%) | 3 (37.5%) | 599 (91.6%) |
| *Missing* | *15 (2.7%)* | *3 (2.5%)* | *8 (1.4%)* | *0 (0%)* | *1 (7.7%)* | *0 (0%)* | *18 (2.7%)* |

Legend: IQR, interquartile range; PCC, post COVID-19 condition; SD, standard deviation.

**Supplementary Figure S1: Sensitivity analysis of post COVID-19 condition and non-recovery and health impairment at 12 months, treating current work ability scores as ordinal outcome.** Current work ability was recategorized into poor (current work ability scores <7), moderate (scores 7–8), and excellent (scores ≥9) work ability for descriptive analyses (panels A–B), and scores from 1–10 were used for multivariable ordinal logistic regression models (panel C). Regression models were adjusted for sex, age, education level, baseline EuroQol visual analog scale (EQ-VAS), comorbidity count, history of psychiatric diagnosis, and hospitalization at acute infection. Separate models were estimated for the two definitions based on COVID-19 related symptoms (PCC vs. no PCC) and (non-)recovery (severe, moderate or mild health impairment vs. recovery). Legend: CI, confidence interval; OR, odds ratio; PCC, post COVID-19 condition; Ref., reference.

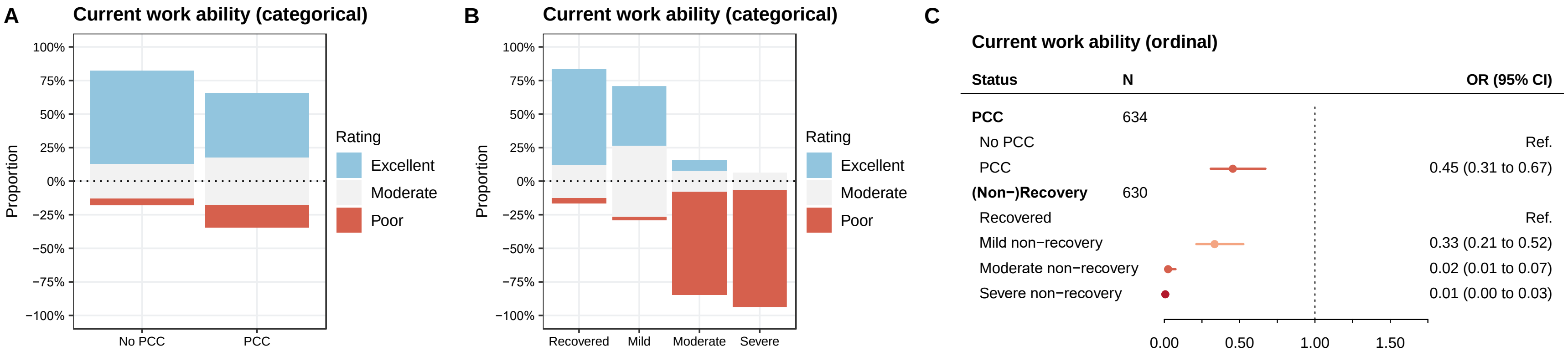

**Supplementary Figure S2: Current work ability stratified by presence of COVID-19 related symptom clusters at 12 months.** Results from descriptive analyses (panel A) and multivariable linear regression analyses (panel B) are presented. Regression models were adjusted for sex, age, education level, baseline EuroQol visual analog scale (EQ-VAS), comorbidity count, history of psychiatric diagnosis, and hospitalization at acute infection. Separate models were estimated for each symptom cluster. Legend: CI, confidence interval; Phys., physical.

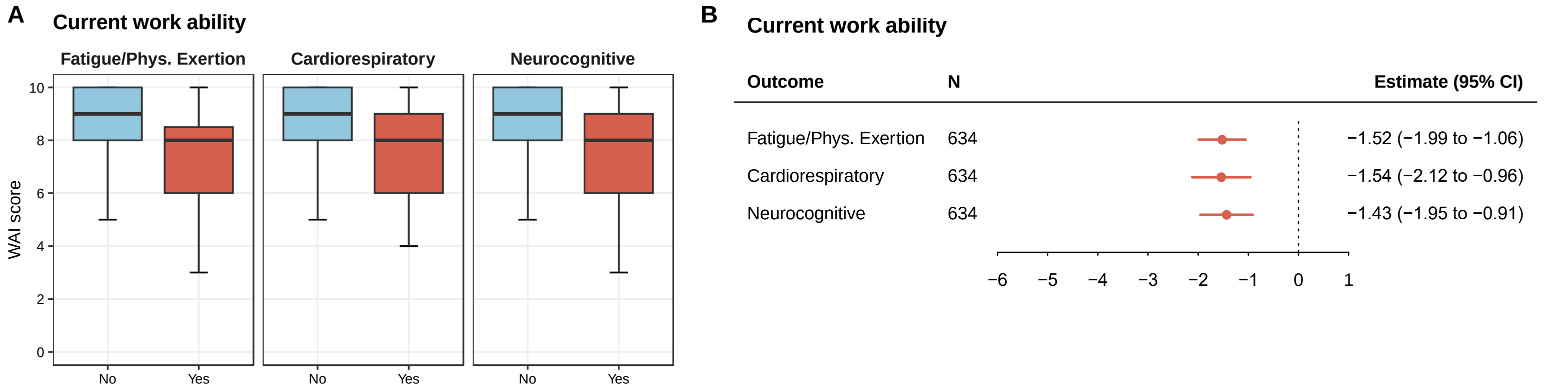

**Supplementary Figure S3: Current work ability stratified by presence of individual COVID-19 related symptom clusters at 12 months.** Results from descriptive analyses (panel A) and multivariable linear regression analyses (panel B) are presented. Regression models were adjusted for sex, age, education level, baseline EuroQol visual analog scale (EQ-VAS), comorbidity count, history of psychiatric diagnosis, and hospitalization at acute infection. Separate models were estimated for each individual symptom. Legend: CI, confidence interval.

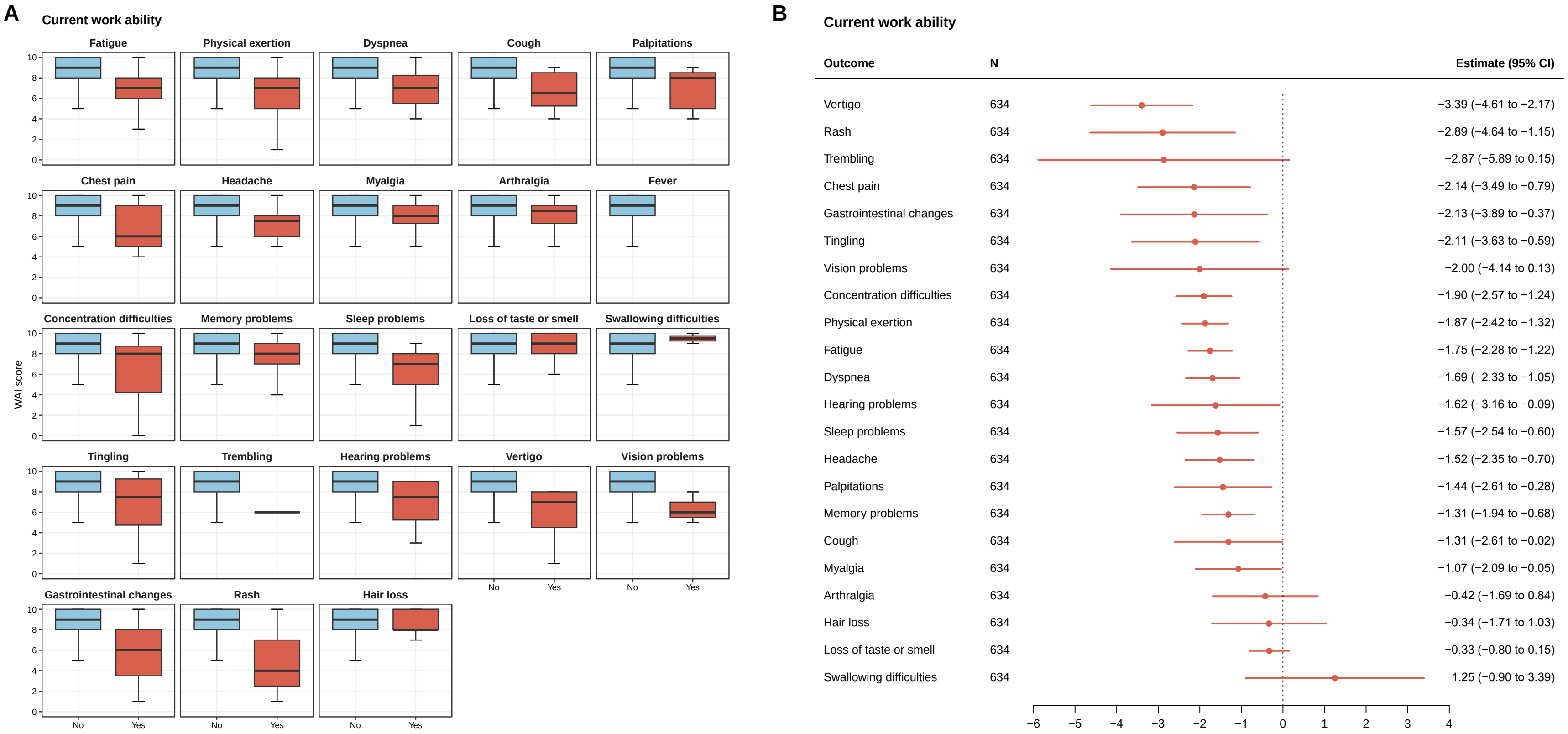

**Supplementary Table S4: Work ability outcomes, stratified by presence of post COVID-19 condition related symptom clusters.**

| | Fatigue/physical exertion | | Cardiorespiratory | | Neurocognitive | |
|---|---|---|---|---|---|---|
| | **Absent (N=620)** | **Present (N=52)** | **Absent (N=642)** | **Present (N=30)** | **Absent (N=635)** | **Present (N=37)** |
| **Current work ability (score)** | | | | | | |
| Mean (SD) | 8.8 (1.6) | 6.8 (2.5) | 8.8 (1.6) | 6.8 (2.7) | 8.8 (1.6) | 6.9 (2.7) |
| Median (IQR) | 9.0 (8.0 to 10.0) | 8.0 (6.0 to 8.5) | 9.0 (8.0 to 10.0) | 8.0 (6.0 to 9.0) | 9.0 (8.0 to 10.0) | 8.0 (6.0 to 9.0) |
| Range | 0 to 10 | 0 to 10 | 0 to 10 | 0 to 10 | 0 to 10 | 0 to 10 |
| *Missing* | *15 (2.4%)* | *1 (1.9%)* | *15 (2.3%)* | *1 (3.3%)* | *16 (2.5%)* | *0 (0%)* |
| **Current work ability (categorical)** | | | | | | |
| Poor (score ≤6) | 31 (5.1%) | 16 (31.4%) | 37 (5.9%) | 10 (34.5%) | 37 (6.0%) | 10 (27.0%) |
| Moderate (score 7-8) | 157 (26.0%) | 22 (43.1%) | 169 (27.0%) | 10 (34.5%) | 163 (26.3%) | 16 (43.2%) |
| Excellent (score ≥9) | 417 (68.9%) | 13 (25.5%) | 421 (67.1%) | 9 (31.0%) | 419 (67.7%) | 11 (29.7%) |
| *Missing* | *15 (2.4%)* | *1 (1.9%)* | *15 (2.3%)* | *1 (3.3%)* | *16 (2.5%)* | *0 (0%)* |
| **Work ability related to physical demands** | | | | | | |
| Very bad | 3 (0.5%) | 2 (4.0%) | 3 (0.5%) | 2 (6.9%) | 4 (0.6%) | 1 (2.8%) |
| Rather bad | 7 (1.2%) | 4 (8.0%) | 10 (1.6%) | 1 (3.4%) | 8 (1.3%) | 3 (8.3%) |
| Moderate | 28 (4.6%) | 17 (34.0%) | 34 (5.4%) | 11 (37.9%) | 37 (6.0%) | 8 (22.2%) |
| Rather good | 138 (22.8%) | 16 (32.0%) | 145 (23.2%) | 9 (31.0%) | 142 (22.9%) | 12 (33.3%) |
| Very good | 429 (70.9%) | 11 (22.0%) | 434 (69.3%) | 6 (20.7%) | 428 (69.1%) | 12 (33.3%) |
| *Missing* | *15 (2.4%)* | *2 (3.8%)* | *16 (2.5%)* | *1 (3.3%)* | *16 (2.5%)* | *1 (2.7%)* |
| **Work ability related to mental demands** | | | | | | |
| Very bad | 3 (0.5%) | 1 (2.0%) | 2 (0.3%) | 2 (6.9%) | 3 (0.5%) | 1 (2.8%) |
| Rather bad | 4 (0.7%) | 3 (6.0%) | 4 (0.6%) | 3 (10.3%) | 4 (0.6%) | 3 (8.3%) |
| Moderate | 55 (9.1%) | 17 (34.0%) | 65 (10.4%) | 7 (24.1%) | 62 (10.0%) | 10 (27.8%) |
| Rather good | 198 (32.8%) | 18 (36.0%) | 205 (32.9%) | 11 (37.9%) | 202 (32.7%) | 14 (38.9%) |

| | | | | | | |
|---|---|---|---|---|---|---|
| Very good | 343 (56.9%) | 11 (22.0%) | 348 (55.8%) | 6 (20.7%) | 346 (56.1%) | 8 (22.2%) |
| *Missing* | *17 (2.7%)* | *2 (3.8%)* | *18 (2.8%)* | *1 (3.3%)* | *18 (2.8%)* | *1 (2.7%)* |
| **Estimated work ability in future (2 years)** | | | | | | |
| Unlikely | 14 (2.3%) | 4 (8.0%) | 15 (2.4%) | 3 (10.7%) | 15 (2.4%) | 3 (8.3%) |
| Not certain | 29 (4.8%) | 8 (16.0%) | 33 (5.3%) | 4 (14.3%) | 32 (5.2%) | 5 (13.9%) |
| Relatively certain | 561 (92.9%) | 38 (76.0%) | 578 (92.3%) | 21 (75.0%) | 571 (92.4%) | 28 (77.8%) |
| *Missing* | *16 (2.6%)* | *2 (3.8%)* | *16 (2.5%)* | *2 (6.7%)* | *17 (2.7%)* | *1 (2.7%)* |

Legend: IQR, interquartile range; PCC, post COVID-19 condition; SD, standard deviation.

**Supplementary Table S5: Results from multivariable regression analyses for work ability outcomes, based on the presence of symptom clusters or specific individual symptoms reported to be related to COVID-19 by participants at 12 months.**

| | Current work ability | | Physical demands | | Mental demands | | Future (2 years) | |
|---|---|---|---|---|---|---|---|---|
| **Symptom** | **Estimate (95% CI)** | **p-value** | **OR (95% CI)** | **p-value** | **OR (95% CI)** | **p-value** | **OR (95% CI)** | **p-value** |
| **Symptom clusters** | | | | | | | | |
| Fatigue/Physical Exertion | -1.52 (-1.99 to -1.06) | <0.001 | 0.15 (0.08 to 0.28) | <0.001 | 0.21 (0.11 to 0.40) | <0.001 | 0.45 (0.18 to 1.10) | 0.079 |
| Cardiorespiratory | -1.54 (-2.12 to -0.96) | <0.001 | 0.17 (0.08 to 0.36) | <0.001 | 0.22 (0.10 to 0.46) | <0.001 | 0.34 (0.12 to 0.92) | 0.033 |
| Neurocognitive | -1.43 (-1.95 to -0.91) | <0.001 | 0.26 (0.13 to 0.50) | <0.001 | 0.23 (0.11 to 0.45) | <0.001 | 0.43 (0.16 to 1.13) | 0.087 |
| **All symptoms** | | | | | | | | |
| Fatigue | -1.75 (-2.28 to -1.22) | <0.001 | 0.13 (0.07 to 0.26) | <0.001 | 0.16 (0.08 to 0.33) | <0.001 | 0.41 (0.15 to 1.10) | 0.076 |
| Physical exertion | -1.87 (-2.42 to -1.32) | <0.001 | 0.13 (0.07 to 0.27) | <0.001 | 0.20 (0.09 to 0.42) | <0.001 | 0.52 (0.18 to 1.50) | 0.227 |
| Dyspnea | -1.69 (-2.33 to -1.05) | <0.001 | 0.17 (0.07 to 0.38) | <0.001 | 0.22 (0.09 to 0.51) | <0.001 | 0.37 (0.12 to 1.09) | 0.071 |
| Cough | -1.31 (-2.61 to -0.02) | 0.047 | 0.27 (0.06 to 1.15) | 0.077 | 0.51 (0.11 to 2.50) | 0.409 | 1.72 (0.15 to 19.99) | 0.665 |
| Palpitations | -1.44 (-2.61 to -0.28) | 0.015 | 0.29 (0.07 to 1.20) | 0.087 | 0.14 (0.03 to 0.63) | 0.011 | 0.10 (0.02 to 0.55) | 0.008 |
| Chest pain | -2.14 (-3.49 to -0.79) | 0.002 | 0.06 (0.01 to 0.38) | 0.003 | 0.03 (0.00 to 0.21) | <0.001 | 0.04 (0.01 to 0.21) | <0.001 |
| Headache | -1.52 (-2.35 to -0.70) | <0.001 | 0.19 (0.07 to 0.56) | 0.003 | 0.21 (0.06 to 0.67) | 0.009 | 0.12 (0.04 to 0.42) | <0.001 |
| Myalgia | -1.07 (-2.09 to -0.05) | 0.040 | 0.25 (0.08 to 0.83) | 0.023 | 0.10 (0.03 to 0.35) | <0.001 | 0.81 (0.09 to 7.17) | 0.849 |
| Arthralgia | -0.42 (-1.69 to 0.84) | 0.509 | 0.16 (0.04 to 0.69) | 0.014 | 0.18 (0.04 to 0.86) | 0.031 | 0.85 (0.08 to 8.52) | 0.889 |
| Concentration difficulties | -1.90 (-2.57 to -1.24) | <0.001 | 0.20 (0.08 to 0.48) | <0.001 | 0.24 (0.09 to 0.60) | 0.002 | 0.54 (0.16 to 1.89) | 0.336 |
| Memory problems | -1.31 (-1.94 to -0.68) | <0.001 | 0.37 (0.16 to 0.82) | 0.015 | 0.35 (0.15 to 0.80) | 0.013 | 0.42 (0.13 to 1.34) | 0.142 |
| Sleep problems | -1.57 (-2.54 to -0.60) | 0.002 | 0.17 (0.05 to 0.57) | 0.004 | 0.13 (0.04 to 0.44) | <0.001 | 0.21 (0.04 to 1.03) | 0.054 |
| Loss of taste or smell | -0.33 (-0.80 to 0.15) | 0.174 | 0.60 (0.32 to 1.13) | 0.117 | 0.59 (0.32 to 1.08) | 0.086 | 0.45 (0.17 to 1.16) | 0.097 |
| Swallowing difficulties | 1.25 (-0.90 to 3.39) | 0.253 | n.e. | | n.e. | | n.e. | |
| Tingling | -2.11 (-3.63 to -0.59) | 0.007 | 0.06 (0.01 to 0.60) | 0.017 | 0.05 (0.01 to 0.43) | 0.006 | 0.13 (0.02 to 0.95) | 0.045 |
| Trembling | -2.87 (-5.89 to 0.15) | 0.063 | 0.00 (0.00 to 0.00) | <0.001 | 0.00 (0.00 to 0.00) | <0.001 | 0.00 (0.00 to 0.00) | <0.001 |

| | | | | | | | | |
|---|---|---|---|---|---|---|---|---|
| Hearing problems | -1.62 (-3.16 to -0.09) | 0.039 | 0.18 (0.02 to 1.89) | 0.153 | 0.24 (0.03 to 1.82) | 0.168 | 0.17 (0.02 to 1.72) | 0.134 |
| Vertigo | -3.39 (-4.61 to -2.17) | <0.001 | 0.04 (0.01 to 0.17) | <0.001 | 0.02 (0.00 to 0.12) | <0.001 | 0.20 (0.03 to 1.14) | 0.070 |
| Vision problems | -2.00 (-4.14 to 0.13) | 0.066 | 0.01 (0.00 to 0.23) | 0.003 | 0.00 (0.00 to 0.04) | <0.001 | 0.01 (0.00 to 0.14) | <0.001 |
| Gastrointestinal changes | -2.13 (-3.89 to -0.37) | 0.018 | 0.01 (0.00 to 0.16) | <0.001 | 0.03 (0.00 to 0.42) | 0.010 | 0.07 (0.01 to 0.56) | 0.013 |
| Rash | -2.89 (-4.64 to -1.15) | 0.001 | 0.07 (0.01 to 0.48) | 0.007 | 0.07 (0.01 to 0.54) | 0.011 | 0.14 (0.02 to 1.03) | 0.053 |
| Hair loss | -0.34 (-1.71 to 1.03) | 0.629 | 0.25 (0.05 to 1.16) | 0.076 | 0.32 (0.07 to 1.50) | 0.148 | 0.08 (0.01 to 0.54) | 0.010 |

Legend: CI, confidence interval; n.e., not estimable; OR, odds ratio. Models are multivariable linear regression models (current work ability) or multivariable ordinal logistic regression models (physical demands, mental demands, future (2 years)) adjusted for age, sex, education status, baseline EuroQol visual analog scale (EQ-VAS), comorbidity count, history of psychiatric diagnosis, and hospitalization due to COVID-19. Reference levels are absence of the respective symptom (cluster).

**Supplementary Figure S4: Current work ability stratified by presence of health problems on any of the standardized health assessments (EQ-5D-5L overall and sub-domains, Fatigue Assessment Scale (FAS), 21-item Depression, Anxiety and Stress scale (DASS-21), modified Medical Research Council (mMRC) dyspnea scale) at 12 months.** Results from descriptive analyses (panel A) and multivariable linear regression analyses (panel B) are presented. Regression models were adjusted for sex, age, education level, baseline EuroQol visual analog scale (EQ-VAS), comorbidity count, history of psychiatric diagnosis, and hospitalization at acute infection. Separate models were estimated for each individual outcome. Legend: CI, confidence interval.

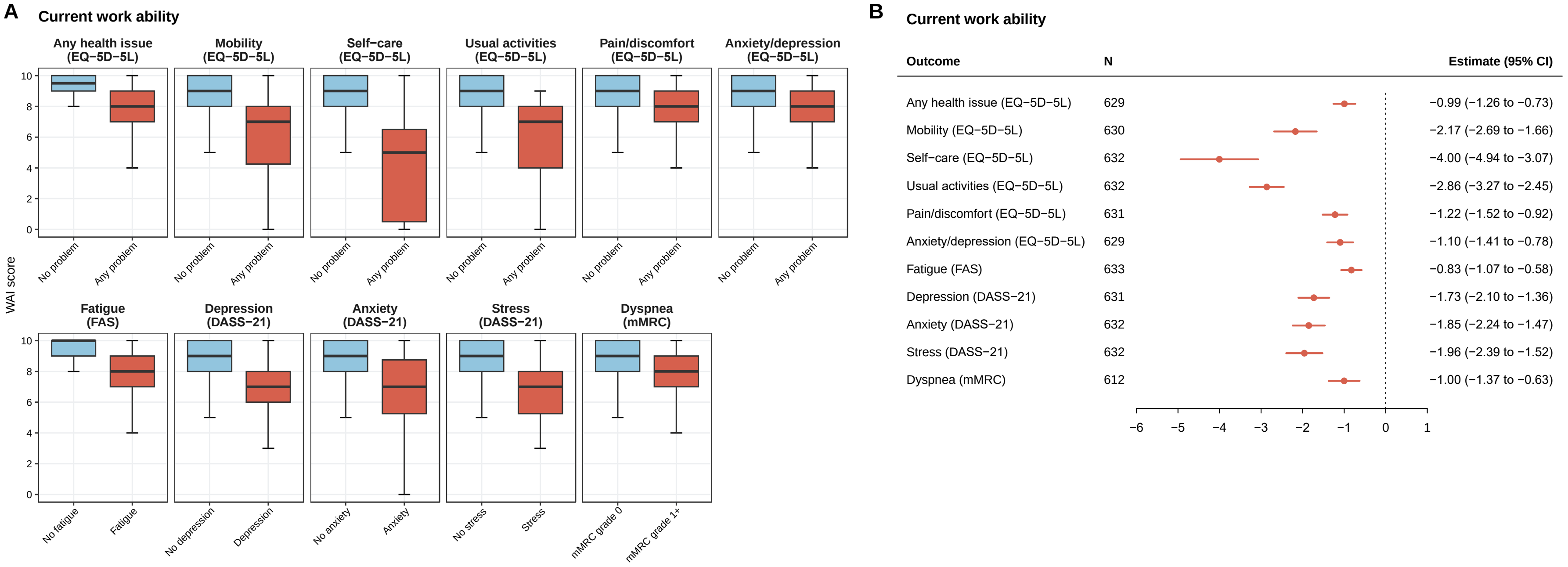

**Supplementary Table S6: Work ability outcomes, stratified by presence of problems on the EQ-5D-5L (overall and subdomains).**

| | **Any health issue (EQ-5D-5L)** | | **Mobility (EQ-5D-5L)** | | **Self-care (EQ-5D-5L)** | | **Usual activities (EQ-5D-5L)** | | **Pain or discomfort (EQ-5D-5L)** | | **Anxiety or depression (EQ-5D-5L)** | |
|---|---|---|---|---|---|---|---|---|---|---|---|---|
| | **No problem (N=431)** | **Any problem (N=222)** | **No problem (N=611)** | **Any problem (N=43)** | **No problem (N=645)** | **Any problem (N=11)** | **No problem (N=595)** | **Any problem (N=61)** | **No problem (N=518)** | **Any problem (N=137)** | **No problem (N=519)** | **Any problem (N=134)** |
| **Current work ability (score)** | | | | | | | | | | | | |
| Mean (SD) | 9.2 (1.2) | 7.8 (2.1) | 8.9 (1.4) | 6.1 (2.8) | 8.8 (1.5) | 4.2 (3.5) | 9.0 (1.2) | 5.8 (2.7) | 9.0 (1.3) | 7.5 (2.4) | 9.0 (1.3) | 7.5 (2.3) |
| Median (IQR) | 9.5 (9.0 to 10.0) | 8.0 (7.0 to 9.0) | 9.0 (8.0 to 10.0) | 7.0 (4.2 to 8.0) | 9.0 (8.0 to 10.0) | 5.0 (0.5 to 6.5) | 9.0 (8.0 to 10.0) | 7.0 (4.0 to 8.0) | 9.0 (8.0 to 10.0) | 8.0 (7.0 to 9.0) | 9.0 (8.0 to 10.0) | 8.0 (7.0 to 9.0) |
| Range | 0 to 10 | 0 to 10 | 0 to 10 | 0 to 10 | 0 to 10 | 0 to 10 | 0 to 10 | 0 to 9 | 0 to 10 | 0 to 10 | 0 to 10 | 0 to 10 |
| *Missing* | *3 (0.7%)* | *1 (0.5%)* | *3 (0.5%)* | *1 (2.3%)* | *4 (0.6%)* | *0 (0%)* | *4 (0.7%)* | *0 (0%)* | *3 (0.6%)* | *1 (0.7%)* | *4 (0.8%)* | *0 (0%)* |
| **Current work ability (categorical)** | | | | | | | | | | | | |
| Poor (score ≤6) | 9 (2.1%) | 36 (16.3%) | 28 (4.6%) | 17 (40.5%) | 38 (5.9%) | 8 (72.7%) | 17 (2.9%) | 29 (47.5%) | 17 (3.3%) | 29 (21.3%) | 19 (3.7%) | 27 (20.1%) |
| Moderate (score 7-8) | 84 (19.6%) | 95 (43.0%) | 161 (26.5%) | 18 (42.9%) | 177 (27.6%) | 2 (18.2%) | 153 (25.9%) | 26 (42.6%) | 123 (23.9%) | 56 (41.2%) | 118 (22.9%) | 59 (44.0%) |
| Excellent (score ≥9) | 335 (78.3%) | 90 (40.7%) | 419 (68.9%) | 7 (16.7%) | 426 (66.5%) | 1 (9.1%) | 421 (71.2%) | 6 (9.8%) | 375 (72.8%) | 51 (37.5%) | 378 (73.4%) | 48 (35.8%) |
| *Missing* | *3 (0.7%)* | *1 (0.5%)* | *3 (0.5%)* | *1 (2.3%)* | *4 (0.6%)* | *0 (0%)* | *4 (0.7%)* | *0 (0%)* | *3 (0.6%)* | *1 (0.7%)* | *4 (0.8%)* | *0 (0%)* |
| **Work ability related to physical demands** | | | | | | | | | | | | |

| | | | | | | | | | | | | |
|---|---|---|---|---|---|---|---|---|---|---|---|---|
| Very bad | 0 (0.0%) | 5 (2.3%) | 2 (0.3%) | 3 (7.1%) | 2 (0.3%) | 3 (27.3%) | 1 (0.2%) | 4 (6.7%) | 0 (0.0%) | 5 (3.7%) | 0 (0.0%) | 5 (3.8%) |
| Rather bad | 2 (0.5%) | 8 (3.6%) | 4 (0.7%) | 6 (14.3%) | 7 (1.1%) | 3 (27.3%) | 2 (0.3%) | 8 (13.3%) | 3 (0.6%) | 7 (5.2%) | 5 (1.0%) | 5 (3.8%) |
| Moderate | 7 (1.6%) | 38 (17.3%) | 35 (5.8%) | 10 (23.8%) | 44 (6.9%) | 1 (9.1%) | 25 (4.2%) | 20 (33.3%) | 14 (2.7%) | 31 (23.0%) | 20 (3.9%) | 25 (18.8%) |
| Rather good | 75 (17.5%) | 78 (35.5%) | 135 (22.2%) | 18 (42.9%) | 151 (23.6%) | 3 (27.3%) | 132 (22.3%) | 22 (36.7%) | 106 (20.6%) | 48 (35.6%) | 110 (21.4%) | 42 (31.6%) |
| Very good | 344 (80.4%) | 91 (41.4%) | 431 (71.0%) | 5 (11.9%) | 436 (68.1%) | 1 (9.1%) | 431 (72.9%) | 6 (10.0%) | 392 (76.1%) | 44 (32.6%) | 380 (73.8%) | 56 (42.1%) |
| *Missing* | *3 (0.7%)* | *2 (0.9%)* | *4 (0.7%)* | *1 (2.3%)* | *5 (0.8%)* | *0 (0%)* | *4 (0.7%)* | *1 (1.6%)* | *3 (0.6%)* | *2 (1.5%)* | *4 (0.8%)* | *1 (0.7%)* |
| **Work ability related to mental demands** | | | | | | | | | | | | |
| Very bad | 0 (0.0%) | 4 (1.8%) | 3 (0.5%) | 1 (2.4%) | 3 (0.5%) | 1 (9.1%) | 1 (0.2%) | 3 (5.0%) | 1 (0.2%) | 3 (2.2%) | 0 (0.0%) | 4 (3.0%) |
| Rather bad | 0 (0.0%) | 6 (2.7%) | 4 (0.7%) | 2 (4.8%) | 5 (0.8%) | 1 (9.1%) | 2 (0.3%) | 4 (6.7%) | 2 (0.4%) | 4 (3.0%) | 0 (0.0%) | 6 (4.5%) |
| Moderate | 21 (4.9%) | 51 (23.2%) | 55 (9.1%) | 17 (40.5%) | 70 (11.0%) | 2 (18.2%) | 43 (7.3%) | 29 (48.3%) | 42 (8.2%) | 30 (22.2%) | 28 (5.5%) | 43 (32.3%) |
| Rather good | 111 (26.1%) | 103 (46.8%) | 202 (33.4%) | 12 (28.6%) | 209 (32.8%) | 6 (54.5%) | 200 (34.0%) | 15 (25.0%) | 157 (30.6%) | 58 (43.0%) | 150 (29.2%) | 65 (48.9%) |
| Very good | 294 (69.0%) | 56 (25.5%) | 341 (56.4%) | 10 (23.8%) | 351 (55.0%) | 1 (9.1%) | 343 (58.2%) | 9 (15.0%) | 311 (60.6%) | 40 (29.6%) | 335 (65.3%) | 15 (11.3%) |
| *Missing* | *5 (1.2%)* | *2 (0.9%)* | *6 (1.0%)* | *1 (2.3%)* | *7 (1.1%)* | *0 (0%)* | *6 (1.0%)* | *1 (1.6%)* | *5 (1.0%)* | *2 (1.5%)* | *6 (1.2%)* | *1 (0.7%)* |

| | | | | | | | | | | | | |
|---|---|---|---|---|---|---|---|---|---|---|---|---|
| **Estimated work ability in future (2 years)** | | | | | | | | | | | | |
| Unlikely | 8 (1.9%) | 10 (4.6%) | 15 (2.5%) | 3 (7.1%) | 16 (2.5%) | 2 (18.2%) | 12 (2.0%) | 6 (10.0%) | 9 (1.7%) | 9 (6.7%) | 12 (2.3%) | 6 (4.5%) |
| Not certain | 13 (3.0%) | 23 (10.5%) | 26 (4.3%) | 10 (23.8%) | 33 (5.2%) | 3 (27.3%) | 22 (3.7%) | 14 (23.3%) | 19 (3.7%) | 17 (12.7%) | 21 (4.1%) | 15 (11.4%) |
| Relatively certain | 407 (95.1%) | 186 (84.9%) | 565 (93.2%) | 29 (69.0%) | 590 (92.3%) | 6 (54.5%) | 556 (94.2%) | 40 (66.7%) | 487 (94.6%) | 108 (80.6%) | 482 (93.6%) | 111 (84.1%) |
| *Missing* | *3 (0.7%)* | *3 (1.4%)* | *5 (0.8%)* | *1 (2.3%)* | *6 (0.9%)* | *0 (0%)* | *5 (0.8%)* | *1 (1.6%)* | *3 (0.6%)* | *3 (2.2%)* | *4 (0.8%)* | *2 (1.5%)* |

Legend: IQR, interquartile range; EQ-5D-5L, EuroQol 5-dimension 5-level scale; SD, standard deviation.

**Supplementary Table S7: Work ability outcomes, stratified by presence of problems on any of the standardized health assessments (FAS, DASS-21, mMRC dyspnea scale).**

| | **Fatigue (FAS)** | | **Depression (DASS-21)** | | **Anxiety (DASS-21)** | | **Stress (DASS-21)** | | **Dyspnea (mMRC)** | |
|---|---|---|---|---|---|---|---|---|---|---|
| | **No fatigue (N=406)** | **Fatigue (N=252)** | **No depression (N=577)** | **Depression (N=78)** | **No anxiety (N=586)** | **Anxiety (N=70)** | **No stress (N=602)** | **Stress (N=54)** | **mMRC grade 0 (N=541)** | **mMRC grade ≥1 (N=93)** |
| **Current work ability (score)** | | | | | | | | | | |
| Mean (SD) | 9.1 (1.4) | 8.0 (2.0) | 9.0 (1.4) | 6.7 (2.5) | 8.9 (1.4) | 6.7 (2.7) | 8.9 (1.5) | 6.5 (2.3) | 8.9 (1.5) | 7.3 (2.4) |
| Median (IQR) | 10.0 (9.0 to 10.0) | 8.0 (7.0 to 9.0) | 9.0 (8.0 to 10.0) | 7.0 (6.0 to 8.0) | 9.0 (8.0 to 10.0) | 7.0 (5.2 to 8.8) | 9.0 (8.0 to 10.0) | 7.0 (5.2 to 8.0) | 9.0 (8.0 to 10.0) | 8.0 (7.0 to 9.0) |
| Range | 0 to 10 | 0 to 10 | 0 to 10 | 0 to 10 | 0 to 10 | 0 to 10 | 0 to 10 | 0 to 10 | 0 to 10 | 0 to 10 |
| *Missing* | *4 (1.0%)* | *0 (0%)* | *3 (0.5%)* | *0 (0%)* | *3 (0.5%)* | *0 (0%)* | *3 (0.5%)* | *0 (0%)* | *2 (0.4%)* | *1 (1.1%)* |
| **Current work ability (categorical)** | | | | | | | | | | |
| Poor (score ≤6) | 13 (3.2%) | 34 (13.5%) | 23 (4.0%) | 24 (30.8%) | 25 (4.3%) | 22 (31.4%) | 26 (4.3%) | 21 (38.9%) | 24 (4.5%) | 22 (23.9%) |
| Moderate (score 7-8) | 78 (19.4%) | 101 (40.1%) | 140 (24.4%) | 36 (46.2%) | 148 (25.4%) | 30 (42.9%) | 153 (25.5%) | 24 (44.4%) | 138 (25.6%) | 37 (40.2%) |
| Excellent (score ≥9) | 311 (77.4%) | 117 (46.4%) | 411 (71.6%) | 18 (23.1%) | 410 (70.3%) | 18 (25.7%) | 420 (70.1%) | 9 (16.7%) | 377 (69.9%) | 33 (35.9%) |
| *Missing* | *4 (1.0%)* | *0 (0%)* | *3 (0.5%)* | *0 (0%)* | *3 (0.5%)* | *0 (0%)* | *3 (0.5%)* | *0 (0%)* | *2 (0.4%)* | *1 (1.1%)* |
| **Work ability related to physical demands** | | | | | | | | | | |
| Very bad | 1 (0.2%) | 4 (1.6%) | 1 (0.2%) | 4 (5.2%) | 1 (0.2%) | 4 (5.7%) | 3 (0.5%) | 2 (3.8%) | 1 (0.2%) | 4 (4.3%) |
| Rather bad | 5 (1.2%) | 6 (2.4%) | 6 (1.0%) | 5 (6.5%) | 6 (1.0%) | 5 (7.1%) | 5 (0.8%) | 6 (11.3%) | 4 (0.7%) | 7 (7.6%) |
| Moderate | 10 (2.5%) | 35 (14.1%) | 29 (5.1%) | 16 (20.8%) | 30 (5.2%) | 15 (21.4%) | 32 (5.3%) | 13 (24.5%) | 21 (3.9%) | 23 (25.0%) |

| | | | | | | | | | | |
|---|---|---|---|---|---|---|---|---|---|---|
| Rather good | 77 (19.1%) | 77 (30.9%) | 122 (21.3%) | 30 (39.0%) | 125 (21.5%) | 27 (38.6%) | 133 (22.2%) | 20 (37.7%) | 116 (21.6%) | 31 (33.7%) |
| Very good | 311 (77.0%) | 127 (51.0%) | 416 (72.5%) | 22 (28.6%) | 420 (72.2%) | 19 (27.1%) | 426 (71.1%) | 12 (22.6%) | 396 (73.6%) | 27 (29.3%) |
| *Missing* | *2 (0.5%)* | *3 (1.2%)* | *3 (0.5%)* | *1 (1.3%)* | *4 (0.7%)* | *0 (0%)* | *3 (0.5%)* | *1 (1.9%)* | *3 (0.6%)* | *1 (1.1%)* |
| **Work ability related to mental demands** | | | | | | | | | | |
| Very bad | 0 (0.0%) | 4 (1.6%) | 0 (0.0%) | 4 (5.2%) | 0 (0.0%) | 4 (5.7%) | 2 (0.3%) | 2 (3.8%) | 1 (0.2%) | 3 (3.3%) |
| Rather bad | 1 (0.2%) | 6 (2.4%) | 2 (0.3%) | 5 (6.5%) | 3 (0.5%) | 4 (5.7%) | 1 (0.2%) | 6 (11.3%) | 3 (0.6%) | 4 (4.3%) |
| Moderate | 16 (4.0%) | 56 (22.6%) | 38 (6.6%) | 34 (44.2%) | 43 (7.4%) | 29 (41.4%) | 46 (7.7%) | 26 (49.1%) | 47 (8.8%) | 23 (25.0%) |
| Rather good | 103 (25.6%) | 113 (45.6%) | 186 (32.5%) | 28 (36.4%) | 188 (32.4%) | 26 (37.1%) | 199 (33.3%) | 16 (30.2%) | 169 (31.5%) | 38 (41.3%) |
| Very good | 283 (70.2%) | 69 (27.8%) | 346 (60.5%) | 6 (7.8%) | 346 (59.7%) | 7 (10.0%) | 349 (58.5%) | 3 (5.7%) | 316 (59.0%) | 24 (26.1%) |
| *Missing* | *3 (0.7%)* | *4 (1.6%)* | *5 (0.9%)* | *1 (1.3%)* | *6 (1.0%)* | *0 (0%)* | *5 (0.8%)* | *1 (1.9%)* | *5 (0.9%)* | *1 (1.1%)* |
| **Estimated work ability in future (2 years)** | | | | | | | | | | |
| Unlikely | 8 (2.0%) | 10 (4.0%) | 11 (1.9%) | 7 (9.0%) | 12 (2.1%) | 6 (8.7%) | 9 (1.5%) | 9 (17.0%) | 10 (1.9%) | 5 (5.5%) |
| Not certain | 12 (3.0%) | 25 (10.0%) | 23 (4.0%) | 13 (16.7%) | 25 (4.3%) | 11 (15.9%) | 27 (4.5%) | 9 (17.0%) | 24 (4.5%) | 12 (13.2%) |
| Relatively certain | 382 (95.0%) | 215 (86.0%) | 538 (94.1%) | 58 (74.4%) | 545 (93.6%) | 52 (75.4%) | 562 (94.0%) | 35 (66.0%) | 504 (93.7%) | 74 (81.3%) |
| *Missing* | *4 (1.0%)* | *2 (0.8%)* | *5 (0.9%)* | *0 (0%)* | *4 (0.7%)* | *1 (1.4%)* | *4 (0.7%)* | *1 (1.9%)* | *3 (0.6%)* | *2 (2.2%)* |

Legend: IQR, interquartile range; DASS-21, 21-item Depression, Anxiety and Stress Scale; FAS, Fatigue Assessment Scale; mMRC, modified Medical Research Council scale; SD, standard deviation.

**Supplementary Table S8: Results from multivariable regression analyses for work ability outcomes, based on the presence of problems in standardized health assessments (EQ-5D-5L, FAS, DASS-21, mMRC dyspnea scale).**

| | **Current work ability** | | **Physical demands** | | **Mental demands** | | **Future (2 years)** | |
|---|---|---|---|---|---|---|---|---|
| **Outcome** | **Estimate (95% CI)** | **p-value** | **OR (95% CI)** | **p-value** | **OR (95% CI)** | **p-value** | **OR (95% CI)** | **p-value** |
| Any health issue (EQ-5D-5L) | -0.99 (-1.26 to -0.73) | <0.001 | 0.19 (0.13 to 0.28) | <0.001 | 0.19 (0.13 to 0.27) | <0.001 | 0.36 (0.18 to 0.71) | 0.003 |
| Mobility (EQ-5D-5L) | -2.17 (-2.69 to -1.66) | <0.001 | 0.11 (0.06 to 0.21) | <0.001 | 0.25 (0.13 to 0.51) | <0.001 | 0.31 (0.12 to 0.82) | 0.018 |
| Self-care (EQ-5D-5L) | -4.00 (-4.94 to -3.07) | <0.001 | 0.03 (0.01 to 0.12) | <0.001 | 0.16 (0.05 to 0.52) | 0.003 | 0.11 (0.03 to 0.46) | 0.002 |
| Usual activities (EQ-5D-5L) | -2.86 (-3.27 to -2.45) | <0.001 | 0.06 (0.03 to 0.11) | <0.001 | 0.10 (0.05 to 0.20) | <0.001 | 0.17 (0.07 to 0.38) | <0.001 |
| Pain/discomfort (EQ-5D-5L) | -1.22 (-1.52 to -0.92) | <0.001 | 0.16 (0.11 to 0.25) | <0.001 | 0.29 (0.19 to 0.43) | <0.001 | 0.27 (0.13 to 0.52) | <0.001 |
| Anxiety/depression (EQ-5D-5L) | -1.10 (-1.41 to -0.78) | <0.001 | 0.25 (0.17 to 0.39) | <0.001 | 0.10 (0.07 to 0.16) | <0.001 | 0.43 (0.21 to 0.89) | 0.023 |
| Fatigue (FAS) | -0.83 (-1.07 to -0.58) | <0.001 | 0.31 (0.22 to 0.44) | <0.001 | 0.18 (0.13 to 0.26) | <0.001 | 0.39 (0.20 to 0.76) | 0.005 |
| Depression (DASS-21) | -1.73 (-2.10 to -1.36) | <0.001 | 0.16 (0.10 to 0.27) | <0.001 | 0.07 (0.04 to 0.13) | <0.001 | 0.17 (0.08 to 0.36) | <0.001 |
| Anxiety (DASS-21) | -1.85 (-2.24 to -1.47) | <0.001 | 0.15 (0.09 to 0.25) | <0.001 | 0.10 (0.06 to 0.17) | <0.001 | 0.19 (0.09 to 0.41) | <0.001 |
| Stress (DASS-21) | -1.96 (-2.39 to -1.52) | <0.001 | 0.13 (0.07 to 0.23) | <0.001 | 0.06 (0.03 to 0.11) | <0.001 | 0.12 (0.05 to 0.27) | <0.001 |
| Dyspnea (mMRC) | -1.00 (-1.37 to -0.63) | <0.001 | 0.20 (0.12 to 0.32) | <0.001 | 0.29 (0.18 to 0.47) | <0.001 | 0.61 (0.27 to 1.36) | 0.224 |

Legend: CI, confidence interval; DASS-21, 21-item Depression, Anxiety and Stress Scale; EQ-5D-5L, EuroQol 5-dimension 5-level scale; FAS, Fatigue Assessment Scale; mMRC, modified Medical Research Council scale; OR, odds ratio. Models are multivariable linear regression models (current work ability) or multivariable ordinal logistic regression models (work ability related to physical and mental demands, estimated future work ability in 2 years) adjusted for age, sex, education status, baseline EuroQol visual analog scale (EQ-VAS), comorbidity count, history of psychiatric diagnosis, and hospitalization due to COVID-19. Reference levels are absence of the respective outcome (based on respective scale).

**Supplementary Table S9: Work ability outcomes, stratified by sex and presence of COVID-19 related symptoms (post COVID-19 condition) at 12 months.**

| | **Female** | | **Male** | |
|---|---|---|---|---|
| | **No PCC (N=285)** | **PCC (N=79)** | **No PCC (N=267)** | **PCC (N=41)** |
| **Current work ability (score)** | | | | |
| Mean (SD) | 8.8 (1.6) | 7.6 (2.5) | 8.9 (1.5) | 8.3 (1.5) |
| Median (IQR) | 9.0 (8.0 to 10.0) | 8.0 (7.0 to 9.0) | 9.0 (8.0 to 10.0) | 9.0 (7.0 to 9.0) |
| Range | 0 to 10 | 0 to 10 | 0 to 10 | 3 to 10 |
| *Missing* | *6 (2.1%)* | *1 (1.3%)* | *9 (3.4%)* | *0 (0%)* |
| **Current work ability (categorical)** | | | | |
| Poor (score ≤6) | 17 (6.1%) | 17 (21.8%) | 10 (3.9%) | 3 (7.3%) |
| Moderate (score 7-8) | 78 (28.0%) | 26 (33.3%) | 59 (22.9%) | 16 (39.0%) |
| Excellent (score ≥9) | 184 (65.9%) | 35 (44.9%) | 189 (73.3%) | 22 (53.7%) |
| *Missing* | *6 (2.1%)* | *1 (1.3%)* | *9 (3.4%)* | *0 (0%)* |
| **Work ability related to physical demands** | | | | |
| Very bad | 0 (0.0%) | 3 (3.9%) | 2 (0.8%) | 0 (0.0%) |
| Rather bad | 3 (1.1%) | 4 (5.2%) | 3 (1.2%) | 1 (2.4%) |
| Moderate | 16 (5.7%) | 15 (19.5%) | 5 (1.9%) | 9 (22.0%) |
| Rather good | 74 (26.4%) | 25 (32.5%) | 41 (16.0%) | 14 (34.1%) |
| Very good | 187 (66.8%) | 30 (39.0%) | 206 (80.2%) | 17 (41.5%) |
| *Missing* | *5 (1.8%)* | *2 (2.5%)* | *10 (3.7%)* | *0 (0%)* |
| **Work ability related to mental demands** | | | | |
| Very bad | 1 (0.4%) | 2 (2.6%) | 1 (0.4%) | 0 (0.0%) |
| Rather bad | 2 (0.7%) | 5 (6.5%) | 0 (0.0%) | 0 (0.0%) |
| Moderate | 32 (11.4%) | 17 (22.1%) | 17 (6.7%) | 6 (14.6%) |
| Rather good | 96 (34.3%) | 27 (35.1%) | 74 (29.0%) | 19 (46.3%) |

| | | | | |
|---|---|---|---|---|
| Very good | 149 (53.2%) | 26 (33.8%) | 163 (63.9%) | 16 (39.0%) |
| *Missing* | *5 (1.8%)* | *2 (2.5%)* | *12 (4.5%)* | *0 (0%)* |
| **Estimated work ability in future (2 years)** | | | | |
| Unlikely | 9 (3.2%) | 6 (7.9%) | 3 (1.2%) | 0 (0.0%) |
| Not certain | 15 (5.4%) | 8 (10.5%) | 9 (3.5%) | 5 (12.2%) |
| Relatively certain | 256 (91.4%) | 62 (81.6%) | 245 (95.3%) | 36 (87.8%) |
| *Missing* | *5 (1.8%)* | *3 (3.8%)* | *10 (3.7%)* | *0 (0%)* |

Legend: IQR, interquartile range; PCC, post COVID-19 condition; SD, standard deviation.

**Supplementary Table S10: Work ability outcomes, stratified by age group and post COVID-19 condition (defined as presence of COVID-19 related symptoms) at 12 months.**

| | 18-39 years | | 40-64 years | |
|---|---|---|---|---|
| | **No PCC (N=249)** | **PCC (N=33)** | **No PCC (N=303)** | **PCC (N=87)** |
| **Current work ability (score)** | | | | |
| Mean (SD) | 8.8 (1.5) | 8.1 (1.9) | 8.9 (1.6) | 7.8 (2.3) |
| Median (IQR) | 9.0 (8.0 to 10.0) | 9.0 (7.0 to 9.0) | 9.0 (8.0 to 10.0) | 8.0 (7.0 to 9.0) |
| Range | 0 to 10 | 1 to 10 | 0 to 10 | 0 to 10 |
| *Missing* | *5 (2.0%)* | *0 (0%)* | *10 (3.3%)* | *1 (1.1%)* |
| **Current work ability (categorical)** | | | | |
| Poor (score ≤6) | 14 (5.7%) | 4 (12.1%) | 13 (4.4%) | 16 (18.6%) |
| Moderate (score 7-8) | 73 (29.9%) | 12 (36.4%) | 64 (21.8%) | 30 (34.9%) |
| Excellent (score ≥9) | 157 (64.3%) | 17 (51.5%) | 216 (73.7%) | 40 (46.5%) |
| *Missing* | *5 (2.0%)* | *0 (0%)* | *10 (3.3%)* | *1 (1.1%)* |
| **Work ability related to physical demands** | | | | |
| Very bad | 0 (0.0%) | 0 (0.0%) | 2 (0.7%) | 3 (3.5%) |
| Rather bad | 2 (0.8%) | 3 (9.1%) | 4 (1.4%) | 2 (2.4%) |
| Moderate | 8 (3.3%) | 3 (9.1%) | 13 (4.4%) | 21 (24.7%) |
| Rather good | 58 (23.8%) | 8 (24.2%) | 57 (19.5%) | 31 (36.5%) |
| Very good | 176 (72.1%) | 19 (57.6%) | 217 (74.1%) | 28 (32.9%) |
| *Missing* | *5 (2.0%)* | *0 (0%)* | *10 (3.3%)* | *2 (2.3%)* |
| **Work ability related to mental demands** | | | | |
| Very bad | 1 (0.4%) | 0 (0.0%) | 1 (0.3%) | 2 (2.4%) |
| Rather bad | 1 (0.4%) | 3 (9.1%) | 1 (0.3%) | 2 (2.4%) |

| | | | | |
|---|---|---|---|---|
| Moderate | 27 (11.1%) | 4 (12.1%) | 22 (7.6%) | 19 (22.4%) |
| Rather good | 90 (36.9%) | 15 (45.5%) | 80 (27.5%) | 31 (36.5%) |
| Very good | 125 (51.2%) | 11 (33.3%) | 187 (64.3%) | 31 (36.5%) |
| *Missing* | *5 (2.0%)* | *0 (0%)* | *12 (4.0%)* | *2 (2.3%)* |
| **Estimated work ability in future (2 years)** | | | | |
| Unlikely | 6 (2.5%) | 0 (0.0%) | 6 (2.0%) | 6 (7.1%) |
| Not certain | 11 (4.5%) | 4 (12.1%) | 13 (4.4%) | 9 (10.7%) |
| Relatively certain | 227 (93.0%) | 29 (87.9%) | 274 (93.5%) | 69 (82.1%) |
| *Missing* | *5 (2.0%)* | *0 (0%)* | *10 (3.3%)* | *3 (3.4%)* |

Legend: IQR, interquartile range; PCC, post COVID-19 condition; SD, standard deviation.

**Supplementary Table S11: Work ability outcomes, stratified by comorbidity count and post COVID-19 condition (defined as presence of COVID-19 related symptoms) at 12 months.**

| | 0-1 comorbidity* | | ≥2 comorbidities* | |
|---|---|---|---|---|
| | **No PCC (N=533)** | **PCC (N=112)** | **No PCC (N=19)** | **PCC (N=8)** |
| **Current work ability (score)** | | | | |
| Mean (SD) | 8.9 (1.4) | 8.0 (2.0) | 7.4 (3.1) | 5.8 (3.3) |
| Median (IQR) | 9.0 (8.0 to 10.0) | 8.0 (7.0 to 9.0) | 9.0 (7.0 to 9.0) | 6.0 (3.8 to 9.0) |
| Range | 0 to 10 | 0 to 10 | 0 to 10 | 0 to 9 |
| *Missing* | *13 (2.4%)* | *1 (0.9%)* | *2 (10.5%)* | *0 (0%)* |
| **Current work ability (categorical)** | | | | |
| Poor (score ≤6) | 24 (4.6%) | 16 (14.4%) | 3 (17.6%) | 4 (50.0%) |
| Moderate (score 7-8) | 132 (25.4%) | 41 (36.9%) | 5 (29.4%) | 1 (12.5%) |
| Excellent (score ≥9) | 364 (70.0%) | 54 (48.6%) | 9 (52.9%) | 3 (37.5%) |
| *Missing* | *13 (2.4%)* | *1 (0.9%)* | *2 (10.5%)* | *0 (0%)* |
| **Work ability related to physical demands** | | | | |
| Very bad | 1 (0.2%) | 1 (0.9%) | 1 (5.9%) | 2 (25.0%) |
| Rather bad | 4 (0.8%) | 3 (2.7%) | 2 (11.8%) | 2 (25.0%) |
| Moderate | 20 (3.8%) | 22 (20.0%) | 1 (5.9%) | 2 (25.0%) |
| Rather good | 111 (21.3%) | 39 (35.5%) | 4 (23.5%) | 0 (0.0%) |
| Very good | 384 (73.8%) | 45 (40.9%) | 9 (52.9%) | 2 (25.0%) |
| *Missing* | *13 (2.4%)* | *2 (1.8%)* | *2 (10.5%)* | *0 (0%)* |
| **Work ability related to mental demands** | | | | |
| Very bad | 2 (0.4%) | 1 (0.9%) | 0 (0.0%) | 1 (12.5%) |
| Rather bad | 2 (0.4%) | 5 (4.5%) | 0 (0.0%) | 0 (0.0%) |

| | | | | |
|---|---|---|---|---|
| Moderate | 45 (8.7%) | 21 (19.1%) | 4 (23.5%) | 2 (25.0%) |
| Rather good | 163 (31.5%) | 44 (40.0%) | 7 (41.2%) | 2 (25.0%) |
| Very good | 306 (59.1%) | 39 (35.5%) | 6 (35.3%) | 3 (37.5%) |
| *Missing* | *15 (2.8%)* | *2 (1.8%)* | *2 (10.5%)* | *0 (0%)* |
| **Estimated work ability in future (2 years)** | | | | |
| Unlikely | 12 (2.3%) | 4 (3.7%) | 0 (0.0%) | 2 (25.0%) |
| Not certain | 20 (3.8%) | 12 (11.0%) | 4 (23.5%) | 1 (12.5%) |
| Relatively certain | 488 (93.8%) | 93 (85.3%) | 13 (76.5%) | 5 (62.5%) |
| *Missing* | *13 (2.4%)* | *3 (2.7%)* | *2 (10.5%)* | *0 (0%)* |

Legend: IQR, interquartile range; PCC, post COVID-19 condition; SD, standard deviation. * Comorbidities were assessed as any of the following: hypertension, diabetes, cardiovascular disease, chronic respiratory disease, chronic kidney disease, past or present malignancy, or immune suppression.

**Supplementary Table S12: Work ability outcomes, stratified by history of psychiatric diagnosis and post COVID-19 condition (defined as presence of COVID-19 related symptoms) at 12 months.**

| | No history of psychiatric diagnosis | | History of psychiatric diagnosis | |
|---|---|---|---|---|
| | **No PCC (N=472)** | **PCC (N=93)** | **No PCC (N=61)** | **PCC (N=25)** |
| **Current work ability (score)** | | | | |
| Mean (SD) | 9.0 (1.4) | 8.4 (1.6) | 8.2 (2.1) | 6.2 (3.2) |
| Median (IQR) | 9.0 (8.0 to 10.0) | 9.0 (8.0 to 10.0) | 9.0 (8.0 to 10.0) | 8.0 (4.0 to 8.0) |
| Range | 0 to 10 | 3 to 10 | 0 to 10 | 0 to 10 |
| *Missing* | *4 (0.8%)* | *1 (1.1%)* | *0 (0%)* | *0 (0%)* |
| **Current work ability (categorical)** | | | | |
| Poor (score ≤6) | 18 (3.8%) | 10 (10.9%) | 7 (11.5%) | 9 (36.0%) |
| Moderate (score 7-8) | 114 (24.4%) | 31 (33.7%) | 21 (34.4%) | 10 (40.0%) |
| Excellent (score ≥9) | 336 (71.8%) | 51 (55.4%) | 33 (54.1%) | 6 (24.0%) |
| *Missing* | *4 (0.8%)* | *1 (1.1%)* | *0 (0%)* | *0 (0%)* |
| **Work ability related to physical demands** | | | | |
| Very bad | 0 (0.0%) | 1 (1.1%) | 2 (3.3%) | 2 (8.3%) |
| Rather bad | 4 (0.9%) | 2 (2.2%) | 1 (1.6%) | 2 (8.3%) |
| Moderate | 14 (3.0%) | 21 (22.8%) | 6 (9.8%) | 3 (12.5%) |
| Rather good | 101 (21.6%) | 31 (33.7%) | 12 (19.7%) | 7 (29.2%) |
| Very good | 349 (74.6%) | 37 (40.2%) | 40 (65.6%) | 10 (41.7%) |
| *Missing* | *4 (0.8%)* | *1 (1.1%)* | *0 (0%)* | *1 (4.0%)* |
| **Work ability related to mental demands** | | | | |
| Very bad | 1 (0.2%) | 1 (1.1%) | 1 (1.6%) | 1 (4.2%) |
| Rather bad | 1 (0.2%) | 2 (2.2%) | 0 (0.0%) | 3 (12.5%) |

| | | | | |
|---|---|---|---|---|
| Moderate | 32 (6.9%) | 16 (17.4%) | 15 (24.6%) | 7 (29.2%) |
| Rather good | 140 (30.0%) | 39 (42.4%) | 28 (45.9%) | 6 (25.0%) |
| Very good | 292 (62.7%) | 34 (37.0%) | 17 (27.9%) | 7 (29.2%) |
| *Missing* | *6 (1.3%)* | *1 (1.1%)* | *0 (0%)* | *1 (4.0%)* |
| **Estimated work ability in future (2 years)** | | | | |
| Unlikely | 9 (1.9%) | 4 (4.3%) | 3 (4.9%) | 2 (8.3%) |
| Not certain | 17 (3.6%) | 9 (9.8%) | 4 (6.6%) | 3 (12.5%) |
| Relatively certain | 442 (94.4%) | 79 (85.9%) | 54 (88.5%) | 19 (79.2%) |
| *Missing* | *4 (0.8%)* | *1 (1.1%)* | *0 (0%)* | *1 (4.0%)* |

Legend: IQR, interquartile range; PCC, post COVID-19 condition; SD, standard deviation.

**Supplementary Table S13: Work ability outcomes, stratified by history of psychiatric diagnosis as well as new or worsened psychiatric diagnoses during follow-up and post COVID-19 condition (defined as presence of COVID-19 related symptoms) at 12 months.**

| | No history and no new psychiatric diagnosis | | No history with new psychiatric diagnosis | | History with no worsened or new psychiatric diagnosis | | History with worsened or new psychiatric diagnosis | |
|---|---|---|---|---|---|---|---|---|
| | **No PCC (N=463)** | **PCC (N=92)** | **No PCC (N=9)** | **PCC (N=1)** | **No PCC (N=44)** | **PCC (N=20)** | **No PCC (N=16)** | **PCC (N=5)** |
| **Current work ability (score)** | | | | | | | | |
| Mean (SD) | 9.0 (1.3) | 8.4 (1.6) | 8.1 (2.0) | 7.0 (NA) | 8.2 (2.2) | 7.2 (2.4) | 8.1 (1.9) | 2.4 (3.4) |
| Median (IQR) | 9.0 (8.0 to 10.0) | 9.0 (8.0 to 10.0) | 8.0 (7.0 to 10.0) | 7.0 (7.0 to 7.0) | 9.0 (8.0 to 10.0) | 8.0 (6.8 to 9.0) | 8.0 (7.8 to 9.2) | 1.0 (0.0 to 3.0) |
| Range | 0 to 10 | 3 to 10 | 4 to 10 | 7 to 7 | 0 to 10 | 1 to 10 | 3 to 10 | 0 to 8 |
| *Missing* | *4 (0.9%)* | *1 (1.1%)* | *0 (0%)* | *0 (0%)* | *0 (0%)* | *0 (0%)* | *0 (0%)* | *0 (0%)* |
| **Current work ability (categorical)** | | | | | | | | |
| Poor (score ≤6) | 17 (3.7%) | 10 (11.0%) | 1 (11.1%) | 0 (0.0%) | 5 (11.4%) | 5 (25.0%) | 2 (12.5%) | 4 (80.0%) |
| Moderate (score 7-8) | 110 (24.0%) | 30 (33.0%) | 4 (44.4%) | 1 (100.0%) | 14 (31.8%) | 9 (45.0%) | 7 (43.8%) | 1 (20.0%) |
| Excellent (score ≥9) | 332 (72.3%) | 51 (56.0%) | 4 (44.4%) | 0 (0.0%) | 25 (56.8%) | 6 (30.0%) | 7 (43.8%) | 0 (0.0%) |
| *Missing* | *4 (0.9%)* | *1 (1.1%)* | *0 (0%)* | *0 (0%)* | *0 (0%)* | *0 (0%)* | *0 (0%)* | *0 (0%)* |
| **Work ability related to physical demands** | | | | | | | | |
| Very bad | 0 (0.0%) | 1 (1.1%) | 0 (0.0%) | 0 (0.0%) | 2 (4.5%) | 1 (5.0%) | 0 (0.0%) | 1 (25.0%) |
| Rather bad | 4 (0.9%) | 2 (2.2%) | 0 (0.0%) | 0 (0.0%) | 1 (2.3%) | 1 (5.0%) | 0 (0.0%) | 1 (25.0%) |
| Moderate | 13 (2.8%) | 20 (22.0%) | 1 (11.1%) | 1 (100.0%) | 1 (2.3%) | 3 (15.0%) | 5 (31.2%) | 0 (0.0%) |
| Rather good | 99 (21.6%) | 31 (34.1%) | 2 (22.2%) | 0 (0.0%) | 9 (20.5%) | 5 (25.0%) | 2 (12.5%) | 2 (50.0%) |
| Very good | 343 (74.7%) | 37 (40.7%) | 6 (66.7%) | 0 (0.0%) | 31 (70.5%) | 10 (50.0%) | 9 (56.2%) | 0 (0.0%) |
| *Missing* | *4 (0.9%)* | *1 (1.1%)* | *0 (0%)* | *0 (0%)* | *0 (0%)* | *0 (0%)* | *0 (0%)* | *1 (20.0%)* |

| | | | | | | | | |
|---|---|---|---|---|---|---|---|---|
| **Work ability related to mental demands** | | | | | | | | |
| Very bad | 0 (0.0%) | 1 (1.1%) | 1 (11.1%) | 0 (0.0%) | 0 (0.0%) | 0 (0.0%) | 1 (6.2%) | 1 (25.0%) |
| Rather bad | 1 (0.2%) | 2 (2.2%) | 0 (0.0%) | 0 (0.0%) | 0 (0.0%) | 1 (5.0%) | 0 (0.0%) | 2 (50.0%) |
| Moderate | 30 (6.6%) | 15 (16.5%) | 2 (22.2%) | 1 (100.0%) | 7 (15.9%) | 6 (30.0%) | 8 (50.0%) | 1 (25.0%) |
| Rather good | 137 (30.0%) | 39 (42.9%) | 3 (33.3%) | 0 (0.0%) | 23 (52.3%) | 6 (30.0%) | 4 (25.0%) | 0 (0.0%) |
| Very good | 289 (63.2%) | 34 (37.4%) | 3 (33.3%) | 0 (0.0%) | 14 (31.8%) | 7 (35.0%) | 3 (18.8%) | 0 (0.0%) |
| *Missing* | *6 (1.3%)* | *1 (1.1%)* | *0 (0%)* | *0 (0%)* | *0 (0%)* | *0 (0%)* | *0 (0%)* | *1 (20.0%)* |
| **Estimated work ability in future (2 years)** | | | | | | | | |
| Unlikely | 7 (1.5%) | 4 (4.4%) | 2 (22.2%) | 0 (0.0%) | 3 (6.8%) | 0 (0.0%) | 0 (0.0%) | 2 (50.0%) |
| Not certain | 17 (3.7%) | 9 (9.9%) | 0 (0.0%) | 0 (0.0%) | 3 (6.8%) | 2 (10.0%) | 1 (6.2%) | 1 (25.0%) |
| Relatively certain | 435 (94.8%) | 78 (85.7%) | 7 (77.8%) | 1 (100.0%) | 38 (86.4%) | 18 (90.0%) | 15 (93.8%) | 1 (25.0%) |
| *Missing* | *4 (0.9%)* | *1 (1.1%)* | *0 (0%)* | *0 (0%)* | *0 (0%)* | *0 (0%)* | *0 (0%)* | *1 (20.0%)* |

Legend: IQR, interquartile range; PCC, post COVID-19 condition; SD, standard deviation.

**Supplementary Table S14: Work ability outcomes, stratified by occupational changes related or unrelated to post COVID-19 condition.**

| | No occupational change (N=540) | Occupational change unrelated to PCC (N=112) | Occupational change related to PCC (N=7) |
|---|---|---|---|
| **Current work ability (score)** | | | |
| Mean (SD) | 8.9 (1.5) | 8.1 (2.1) | 3.4 (1.9) |
| Median (IQR) | 9.0 (8.0 to 10.0) | 8.5 (7.0 to 10.0) | 4.0 (2.0 to 4.5) |
| Range | 0 to 10 | 0 to 10 | 1 to 6 |
| *Missing* | *1 (0.2%)* | *2 (1.8%)* | *0 (0%)* |
| **Current work ability (categorical)** | | | |
| Poor (score ≤6) | 25 (4.6%) | 15 (13.6%) | 7 (100.0%) |
| Moderate (score 7-8) | 139 (25.8%) | 40 (36.4%) | 0 (0.0%) |
| Excellent (score ≥9) | 375 (69.6%) | 55 (50.0%) | 0 (0.0%) |
| *Missing* | *1 (0.2%)* | *2 (1.8%)* | *0 (0%)* |
| **Work ability related to physical demands** | | | |
| Very bad | 3 (0.6%) | 1 (0.9%) | 1 (14.3%) |
| Rather bad | 5 (0.9%) | 4 (3.6%) | 2 (28.6%) |
| Moderate | 35 (6.5%) | 8 (7.2%) | 2 (28.6%) |
| Rather good | 115 (21.4%) | 37 (33.3%) | 2 (28.6%) |
| Very good | 379 (70.6%) | 61 (55.0%) | 0 (0.0%) |
| *Missing* | *3 (0.6%)* | *1 (0.9%)* | *0 (0%)* |
| **Work ability related to mental demands** | | | |
| Very bad | 1 (0.2%) | 2 (1.8%) | 1 (14.3%) |
| Rather bad | 2 (0.4%) | 2 (1.8%) | 3 (42.9%) |
| Moderate | 50 (9.3%) | 19 (17.1%) | 3 (42.9%) |

| | | | |
|---|---|---|---|
| Rather good | 165 (30.8%) | 51 (45.9%) | 0 (0.0%) |
| Very good | 317 (59.3%) | 37 (33.3%) | 0 (0.0%) |
| *Missing* | *5 (0.9%)* | *1 (0.9%)* | *0 (0%)* |
| **Estimated work ability in future (2 years)** | | | |
| Unlikely | 13 (2.4%) | 3 (2.7%) | 2 (33.3%) |
| Not certain | 21 (3.9%) | 13 (11.8%) | 3 (50.0%) |
| Relatively certain | 504 (93.7%) | 94 (85.5%) | 1 (16.7%) |
| *Missing* | *2 (0.4%)* | *2 (1.8%)* | *1 (14.3%)* |

Legend: IQR, interquartile range; PCC, post COVID-19 condition; SD, standard deviation.
